# Prediction of recessive inheritance for missense variants in human disease

**DOI:** 10.1101/2021.10.25.21265472

**Authors:** Ben O. Petrazzini, Daniel J. Balick, Iain S. Forrest, Judy Cho, Ghislain Rocheleau, Daniel M. Jordan, Ron Do

## Abstract

The prediction of pathogenic human missense variants has improved in recent years, but a more granular level of variant characterization is required. Further axes of information need to be incorporated in order to advance the genotype-to-phenotype map. Recent efforts have developed mode of inheritance prediction tools; however, these lack robust validation and their discrimination performance does not support clinical utility, with evidence of them being fundamentally insensitive to recessive acting diseases. Here, we present MOI-Pred, a three-way variant-level mode of inheritance prediction tool aimed at recessive identification for missense variants. MOI-Pred shows strong ability to discriminate missense variants causing autosomal recessive disease (area under the receiver operating characteristic (AUROC)=0.99 and sensitivity=0.85) in an external validation set. Additionally, we introduce an electronic health record (EHR)-based validation approach using real-world clinical data and show that our recessive predictions are enriched for recessive associations with human diseases, demonstrating utility of our method. Mode of inheritance predictions - pathogenic for autosomal recessive (AR) disease, pathogenic for autosomal dominant (AD) disease, or benign – for all possible missense variants in the human genome are available at https://github.com/rondolab/MOI-Pred/.

## Introduction

Computational methods to predict the effect of coding variants have numerous applications, such as the diagnosis of genetic diseases^1-4^, genetic association studies^5-8^, and drug design^9,10^. Currently available methods perform very well at discriminating pathogenic and benign missense variants, typically reporting accuracy in the range of 58-86%^11-13^. Each prediction uses a unique set of variant characteristics and shows different performances across datasets making ensemble approaches more accurate^14-16^. On the same basis, current guidelines recommend considering multiple prediction tools to inform decision making^13^. While these methods may perform very well, they do not consider granularity of a variant’s effect on disease. The vast majority of these methods make a simple binary prediction: is the variant pathogenic, or is it benign? Some methods make more specific predictions about whether variants cause particular phenotypes^17^, but even these are generally still binary predictions about a single phenotype. The true shape of the genotype-to-phenotype map is much more complex and highly dimensional. A full assessment of a variant’s effect on phenotype would include potentially pleiotropic effects on a variety of different phenotypes, from the molecular level to the systems level, as well as features that modify its genetic impact, such as penetrance and mode of inheritance. Large-scale computational approaches that incorporate different axes of genomic information can potentially be used to inform various aspects of variant function^18^.

Here, we focus on mode of inheritance as the next level of granularity to include in computational prediction of variant effect. The concept behind mode of inheritance is foundational to the field of genetics and in classical medical genetics it is considered one of the most important features to report about a pathogenic variant^13,19,20^. Studies have shown that disease diagnosis can be largely improved by incorporating pedigree information^21-24^. In spite of this, mode of inheritance has practically no role in current variant annotation pipelines. Efforts to resolve mode of inheritance mechanisms have fallen behind the gene discovery rate^25^, limiting the availability of such information in databases of validated clinically relevant variants. Even among databases that do provide mode of inheritance information, most notably Online Mendelian Inheritance in Man (OMIM)^26^, these annotations are present only for a small fraction of 4,417 genes. They are also not necessarily reliable, since they derive almost entirely from anecdotal case reports with small pedigrees, and very few have been replicated across studies. Currently, 35.5% of variants reported as “Pathogenic” or “Likely Pathogenic” in ClinVar^27^ have no annotated mode of inheritance and cannot confidently be assigned one based on existing annotations. While some molecular and evolutionary features are known to be enriched in genes implicated in autosomal recessive (AR) disease^28-33^, these features are not widely used at the variant level to distinguish variants causing AR disease from variants causing autosomal dominant (AD) disease or benign variants. Additionally, there is some evidence that current variant effect prediction methods may be fundamentally insensitive to AR disease^34,35^, reinforcing the need for new methods specifically aimed at predicting variants causing AR disease^36^. Previous efforts at developing such methods underperform binary prediction tools, lack robust validation and have not achieved widespread use in the field^16,37-39^.

Here we present MOI-Pred, a three-way prediction method that labels missense variants as pathogenic for AR disease, pathogenic for AD disease, or benign. The method uses a random forest classifier to combine variant effect estimations with gene-level features that are predictive of AR or AD disease. The resulting predictor identifies pathogenic mutations with performance comparable to state-of-the-art binary prediction methods and distinguishes mode of inheritance at the variant level. Moreover, the tool accurately predicts disease case-control status for the three classes of mutations in an external validation using real-world electronic health record (EHR)-based clinical data. MOI-Pred addresses a shortcoming in current annotation pipelines by accurately predicting mode of inheritance, especially differentiating AR pathogenic variants from benign variants, while simultaneously improving granular predictions of variant effect crucial to achieve clinically relevant levels of accuracy.

## Results

### Clinical variants missing mode of inheritance information

ClinVar does not explicitly annotate mode of inheritance. Instead, this information is extracted from external resources such as OMIM or the Human Gene Mutation Database (HGMD)^40^. These databases provide mainly gene-level information and only for a subset of diseases. Thus, most variants in ClinVar either lack a mode of inheritance annotation entirely or are simply labelled with the annotation of their corresponding gene. Only 4,126 genes in ClinVar have inheritance information, resulting in 37.63% of variants with undetermined mode of inheritance (**Supplementary Table 2**). Out of a total of 307,800 unlabelled variants, 49,745 are Pathogenic (35.51% of all Pathogenic), 119,532 are Benign (41.38% of all Benign), 122,600 have Uncertain significance (35.30% of all Uncertain significance) and 15,923 have Conflicting interpretation of pathogenicity (38.19% of all Conflicting interpretation) (**Supplementary Table 2**).

### Model Training

We collected a training set of 2,481 Recessive and 1,248 Dominant pathogenic missense variants from ExoVar^16^ and 3,729 Benign missense variants from gnomAD^41^, annotated with a wide range of features capturing functional and biological aspects of mode of inheritance. We fitted a random forest model on this training set, using 10-fold cross-validation and 100 different random train-test splits to assess performance. Feature selection was performed independently on each iteration, reducing the number of features to a minimum of 10 and a maximum of 18 (median across 100 models is 13 features (**Supplementary Figure 1**). In total, 19 unique features were selected across all 100 iterations for training, incorporating a range of functional, evolutionary and combined information (**Methods, Supplementary Table 1**).

The prediction models performed well in the Test set, with a mean area under the receiver operator characteristic (AUROC)=0.94/0.96/0.95 (standard deviation (SD)=1.2 × 10^−2^/6.8 × 10^−3^ /1.3 × 10^−2^), sensitivity=0.75/0.76/0.92 (SD=3.8 × 10^−2^/3.0 × 10^−2^/2.8 × 10^−2^), and specificity=0.94/0.95/0.82 (SD=1.3 × 10^−2^/1.4 × 10^−2^/2.2 × 10^−2^) for Recessive/Dominant/Benign classes, respectively (**Figure 2**). This represents good overall performance with similar discrimination power across classes. Benign has higher sensitivity and lower specificity than the other two classes, representing a higher rate of false positives for Benign variants and a higher rate of false negatives for both classes of pathogenic variants.

**Figure 1.**
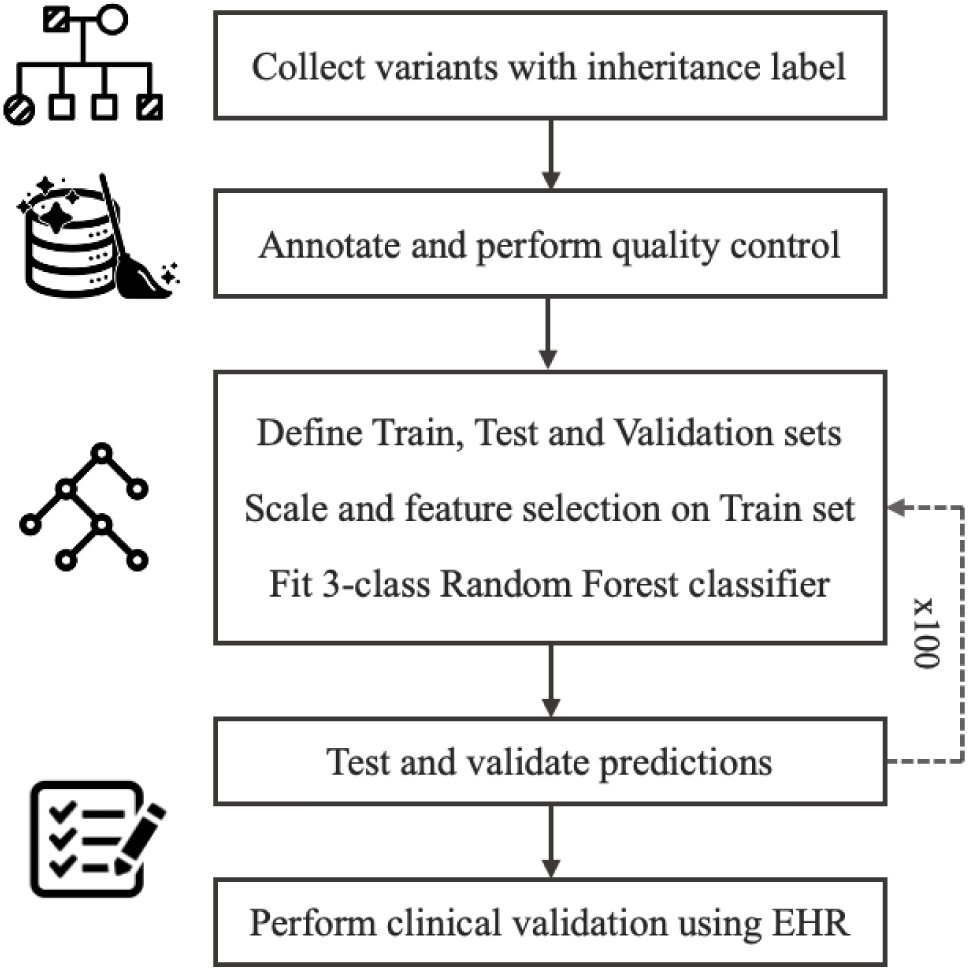
Study design and machine learning workflow. Train and Test sets correspond to the 90% and 10% balanced datasets, built from ExoVar and gnomAD variants, used for training and testing respectively. Validation set corresponds to the balanced dataset, built from ClinVar and GEM variants, used for external validation. EHR corresponds to electronic health records.

**Figure 2.**
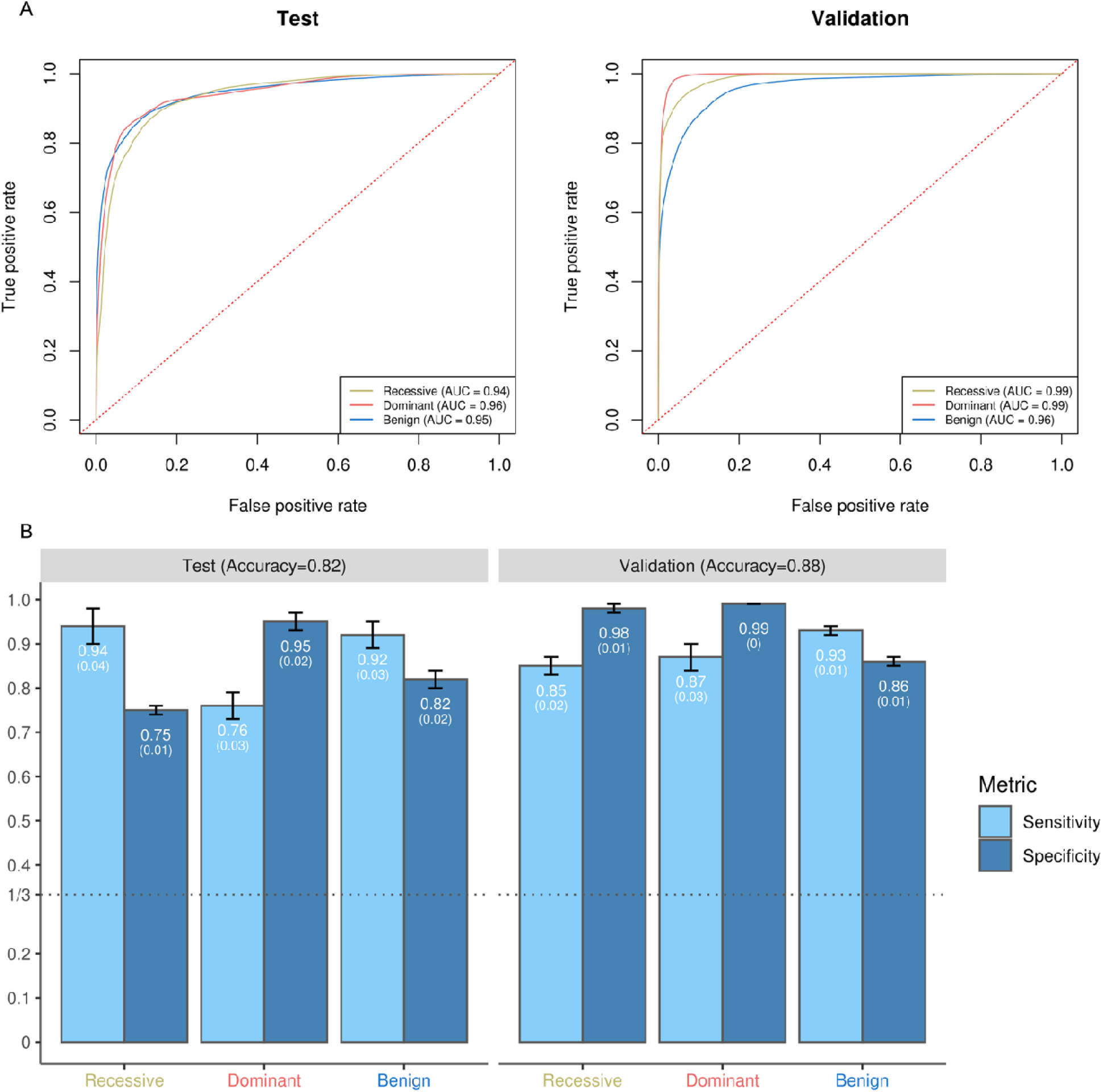
Receiver operator characteristic curves for 3-class mode of inheritance prediction models (A). Bar-plots showing sensitivity and specificity for 3-class mode of inheritance prediction models (B). Test set corresponds to the 10% balanced dataset, built from ExoVar and gnomAD variants, used for testing. Validation set corresponds to the balanced dataset, built from ClinVar and GEM variants, used for external validation. Reported AUC corresponds to the mean across 100 models. Reported Sensitivity and Specificity corresponds to mean (standard deviation) across 100 models. AUC corresponds to area under the receiver operator characteristic curve.

### External Validation

To assess the model’s performance on external data sources, we collected an external Validation set containing 261 Recessive and 255 Dominant pathogenic missense variants from ClinVar and 1,010 Benign missense variants from the Japan Whole Genome Aggregation panel v.1 (GEM)^42^, in addition to the internal Train/Test set. Performance on this external Validation set was similar to performance measured on the Test set, with AUROC=0.99/0.99/0.96 (SD=2.7 × 10^−3^/1.8 × 10^−3^/6.9 × 10^−3^), sensitivity 0.85/0.87/0.93 (SD 1.8 × 10^−2^/2.8 × 10^−2^/1.7 × 10^−2^), and specificity 0.98/0.99/0.86 (SD 7.4 × 10^−3^/4.8 × 10^−3^/1.5 × 10^−2^) for Recessive/Dominant/Benign classes (**Figure 2**). This suggests that the model is not overfitting the data sources used for training and testing (ExoVar and gnomAD), which would be indicated by a significant drop in performance between the internal blind Test set and external validation. Indeed, the model appears to perform better on the external validation set. This may reflect the fact that the ClinVar datasets used for external validation contain more confident annotations and less noise than the ExoVar datasets used for training and testing.

To expand on this observation, we grouped variants by ClinVar review status (level of evidence for pathogenicity) and assessed how the confidence of variant annotations affects the sensitivity of our predictions. We found that sensitivity improved with higher review status: sensitivity for the Recessive/Dominant classes was 0.58/0.49 (SD 0.03/0.00) for 0-star review status, 0.68/0.59 (SD 0.04/0.00) for 1-star review status, and 0.86/0.89 (SD 0.04/0.00) for 2-star or higher review status (**Supplementary Figure 2**). This is as expected if lower-confidence variants are less likely to be truly pathogenic. In this case, an accurate predictor would predict a smaller fraction of 0-star variants to be pathogenic, because the fraction of those variants that are truly pathogenic is smaller.

We also tested the inheritance prediction model on variants unique to a single ancestry group (European American, African American, or Hispanic American) to evaluate whether performance is consistent across ancestries. We found that sensitivity was uniformly high across all three ancestries, with no specific ancestry having substantially higher power (**Supplementary Figure 3, 4 and 5**). We also found that ancestry-specific variants across all three ancestries showed the same trend as the full dataset, with sensitivity improving in higher confidence annotations. This demonstrates that MOI-Pred is not primarily powered to detect variants observed in Europeans, but has similar performance regardless of ancestry.

### Model interpretation

Examining the importance of different features in the model shows the union of functional, evolutionary and combined information that are driving the inheritance prediction. One functional feature (AD.rank with 23.8%), two combined features (MutPred and MCAP with 14.2% and 13.6%, respectively) and two evolutionary features (OE and FATHMM with 11.2% and 11% respectively) carry 73.8% of the models’ weight (**Figure 3. A**).

**Figure 3:**
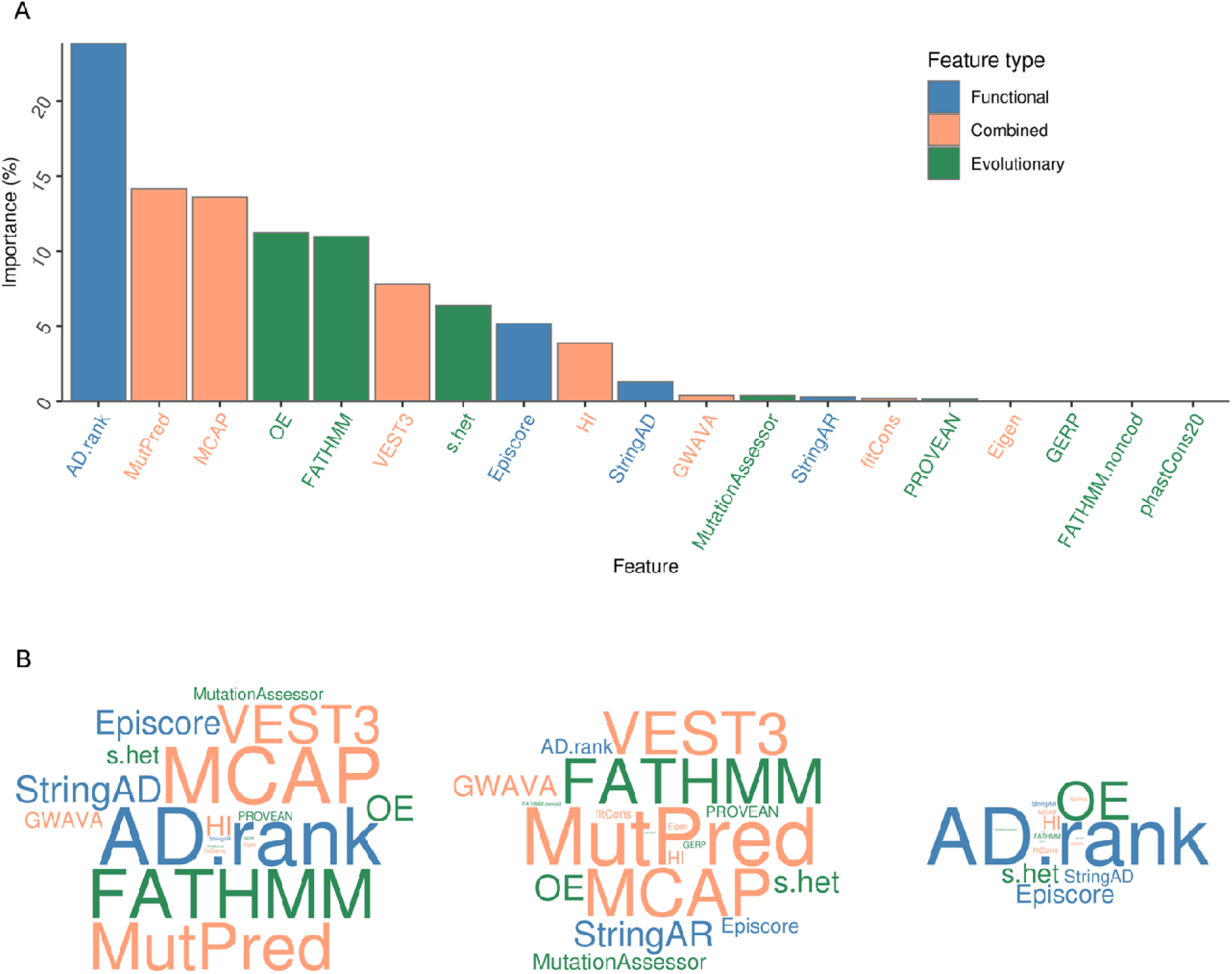
Bar-plot showing feature importance on 3-class mode of inheritance prediction models (A). Word-clouds representing feature importance on 2-class mode of inheritance prediction models (B). Feature importance is reported as the median across 100 models. Word-clouds represent feature importance on Benign-Dominant (left), Benign-Recessive (middle) and Dominant-Recessive (right) models respectively. Exact feature importance values in 2-class prediction models can be found in Supplementary Figure 6-8.

These feature weights represent the overall importance of features to the three-way classifier. To examine which features are important to identify each individual class, we trained three two-way classifiers to distinguish Dominant from Benign, Recessive from Benign, and Dominant from Recessive. Both Benign-Pathogenic binary prediction models (Benign-Dominant and Benign-Recessive) are dominated by features carrying combined functional and evolutionary information, namely MCAP, MutPred and VEST3, in addition to FATHMM which primarily carries evolutionary information (**Figure 3. B, Supplementary Figure 6 and 7**). In contrast, the Dominant-Recessive prediction is mainly driven by gene-level features carrying either functional or evolutionary information like AD.rank and O/E (**Figure 3. B, Supplementary Figure 8**).

### Clinical validation using EHR

To test the performance of the model on real-world clinical data, we collected a total of 1,845,623 variants present in patients from the Bio*Me* biobank^43^. Of these, 56,706 were missense variants present in ClinVar (2,301 Pathogenic, 9,865 Benign, 35,629 Uncertain significance and 8,911 Conflicting interpretation), and 19,134 remain after restricting to 2-star or higher in ClinVar review status (1,047 Pathogenic, 6,303 Benign and 11,784 Uncertain significance) (**Supplementary Table 3**). The model used to predict all variants shows good performance in the Train/Test and Validation sets (**Supplementary Table 4**). For each variant, we marked each patient as positive if the EHR included a diagnosis reported for the variant in ClinVar, and negative otherwise. We then used a Cochran-Mantel-Haenszel (CMH) stratified contingency test to assess the association between homozygous or heterozygous carriers of ClinVar-annotated variants and actual diagnoses, stratified by disease. An association between carrier status and disease status indicates that the variants being tested are, in aggregate, associated with disease with the specified mode of inheritance. By separating variants that receive different predictions from our model, we can test whether our model’s prediction is actually predictive of carrier disease status in a real clinical population.

The contingency table analysis showed that our model is highly predictive, with all ClinVar categories showing associations in the expected directions. We found that a Recessive prediction significantly increases the association between homozygous carrier status and disease status for all ClinVar Pathogenic (OR=4.30 [95% CI=4.07 to 4.55] and OR=1.07 [95% CI=1.04 to 1.09]), Uncertain Significance (OR=5.45 [95% CI=5.13 to 5.77] and OR=0.31 [95% CI=0.30 to 0.32]) and Conflicting Interpretation (OR=4.11 [95% CI=3.94 to 4.28] and OR=1.11 [95% CI=1.09 to 1.13]) annotations. A Dominant prediction significantly increases the association between homozygous or heterozygous carrier status and disease status for ClinVar Pathogenic (Odds ratio (OR)=1.98 for MOI-Pred’s prediction [95% confidence interval (CI)=1.96 to 2.00] and OR=1.56 [95% CI=1.55 to 1.57] for all other variants) and Uncertain Significance (OR=1.40 [95% CI=1.39 to 1.41 and OR=0.87 [95% CI=0.87 to 0.87]) annotations. And as expected, a Benign prediction significantly decreases the association between homozygous or heterozygous carrier status and disease status for ClinVar Pathogenic (OR=1.23 [95% CI=1.22 to 1.24] and OR=2.97 [95% CI=2.94 to 2.99]) and Uncertain Significance (OR=0.87 [95% CI=0.87 to 0.88] and OR=1.02 [95% CI=1.01 to 1.02]) annotations (**Figure 4, Supplementary Table 5-7**). Restricting to ClinVar variants with 2-star or higher review status showed similar results (**Supplementary Figure 9, Supplementary Table 8-10**). Notably, we observed a particularly strong protective association of variants on disease that are not predicted recessive for “Uncertain Significance” variants (**Figure 4C**).

**Figure 4:**
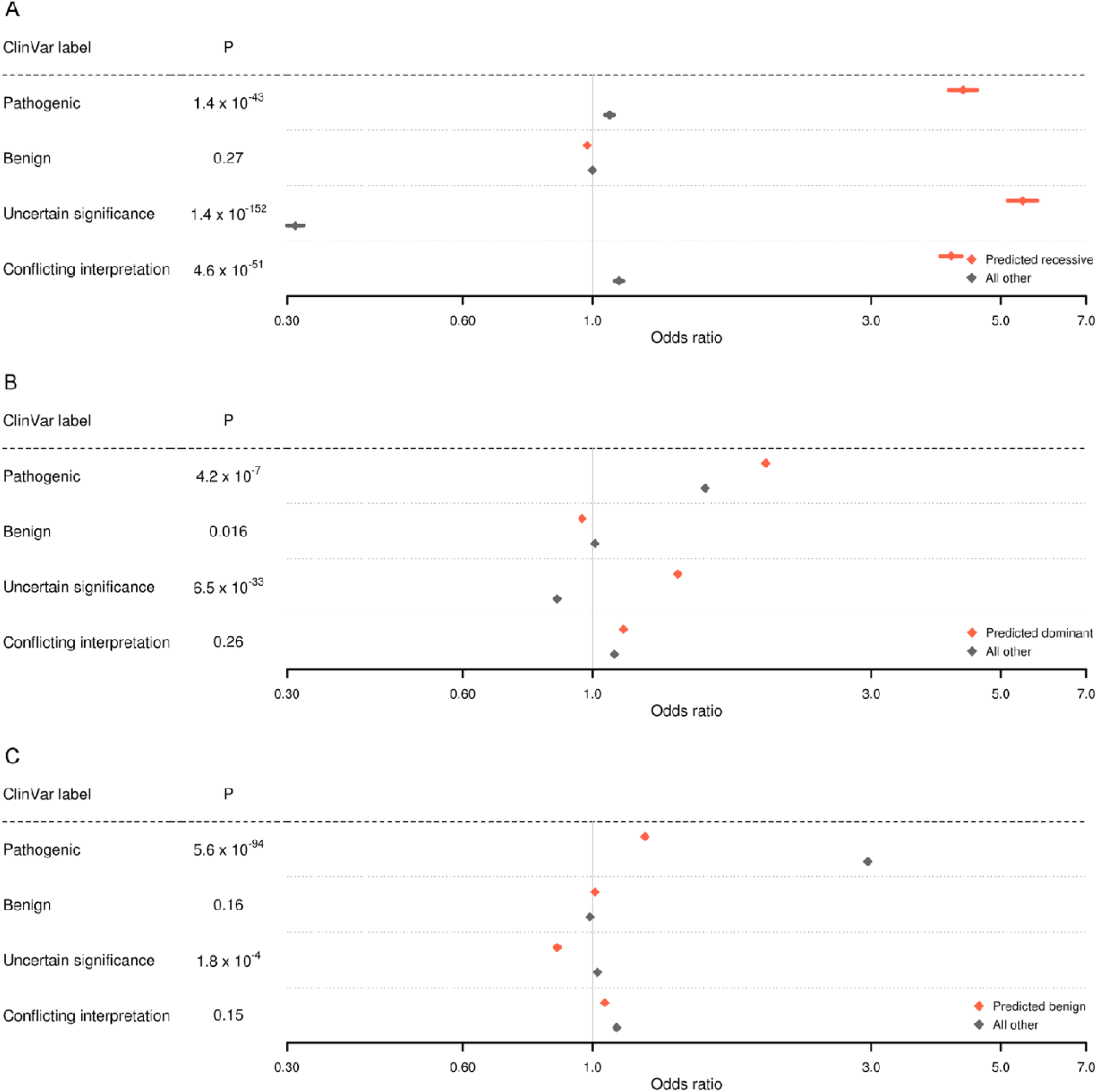
Forest plots showing disease association with variants predicted to be Recessive (A), Dominant (B) and Benign (C). Effect sizes (Odds ratios) and 95% confidence intervals were obtained for individual ancestries using a Cochran-Mantel-Haenszel (CMH) test. The reported effect sizes correspond to an inverse variance meta-analysis across ancestries. P values for heterogeneity between Odds ratios are derived from a Q-test Test set.

To compare MOI-Pred with a previously developed mode of inheritance prediction tool (MAPPIN), we also performed the same EHR-based clinical validation analyses using MAPPIN predictions (**Supplementary Figure 10, Supplementary Table 11-13**). MAPPIN shows weaker enrichment for recessive association with disease of ClinVar Pathogenic predicted recessive (OR=3.61 [95% CI=3.41 to 3.80] for MAPPIN vs. OR=4.31 [95% CI=4.07 to 4.55] for MOI-Pred) and no enrichment for recessive association with disease of ClinVar Uncertain Significance and Conflicting Interpretation variants (OR=0.35 [95% CI=0.33 to 0.36] for MAPPIN vs. OR=5.45 [95% CI=5.13 to 5.77] for MOI-Pred and OR=1.15 [95% CI=1.11 to 1.18] for MAPPIN vs. OR=4.11 [95% CI=3.94 to 4.28] for MOI-Pred, respectively) (**Supplementary Figure 10, Supplementary Table 11**). As expected, MAPPIN shows similar or stronger enrichment for dominant association with disease for various ClinVar classes (**Supplementary Figure 10, Supplementary Table 12**).

### Discovery of individual variants

To test the utility of MOI-Pred for clinical assessment of individual variants, we performed single variant association tests in the Bio*Me* biobank and identified 18 variants showing significant associations with a single phenotype (p-value corrected for 455 recessive association tests=1.09 × 10^−4^; p-value corrected for 455 recessive association tests for 6,382 dominant association tests=7.83 × 10^−6^) (**Table 1 and Supplementary Table 14-17**). Three variants were found to have significant recessive associations with disease. Interestingly, none of these are labelled as Pathogenic in ClinVar as two are labelled Benign and one is labelled Conflicting Interpretation of Pathogenicity. Moreover, 15 variants showed dominant association with disease, 12 of which do not correspond with their clinical annotation using ClinVar/OMIM, showing the potential utility of MOI-Pred for discovery of novel associations with disease (**Table 1**).

**Table 1:**
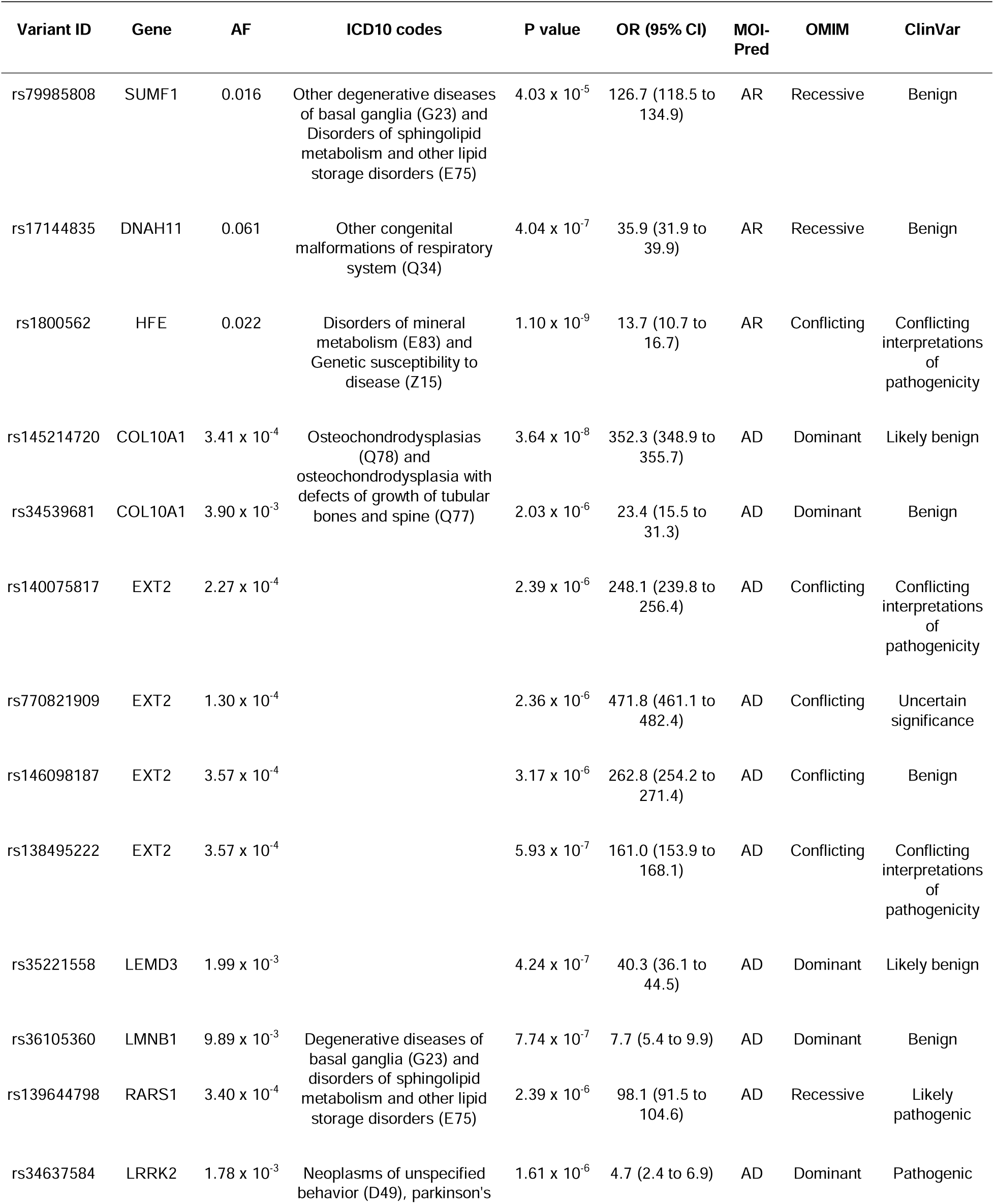
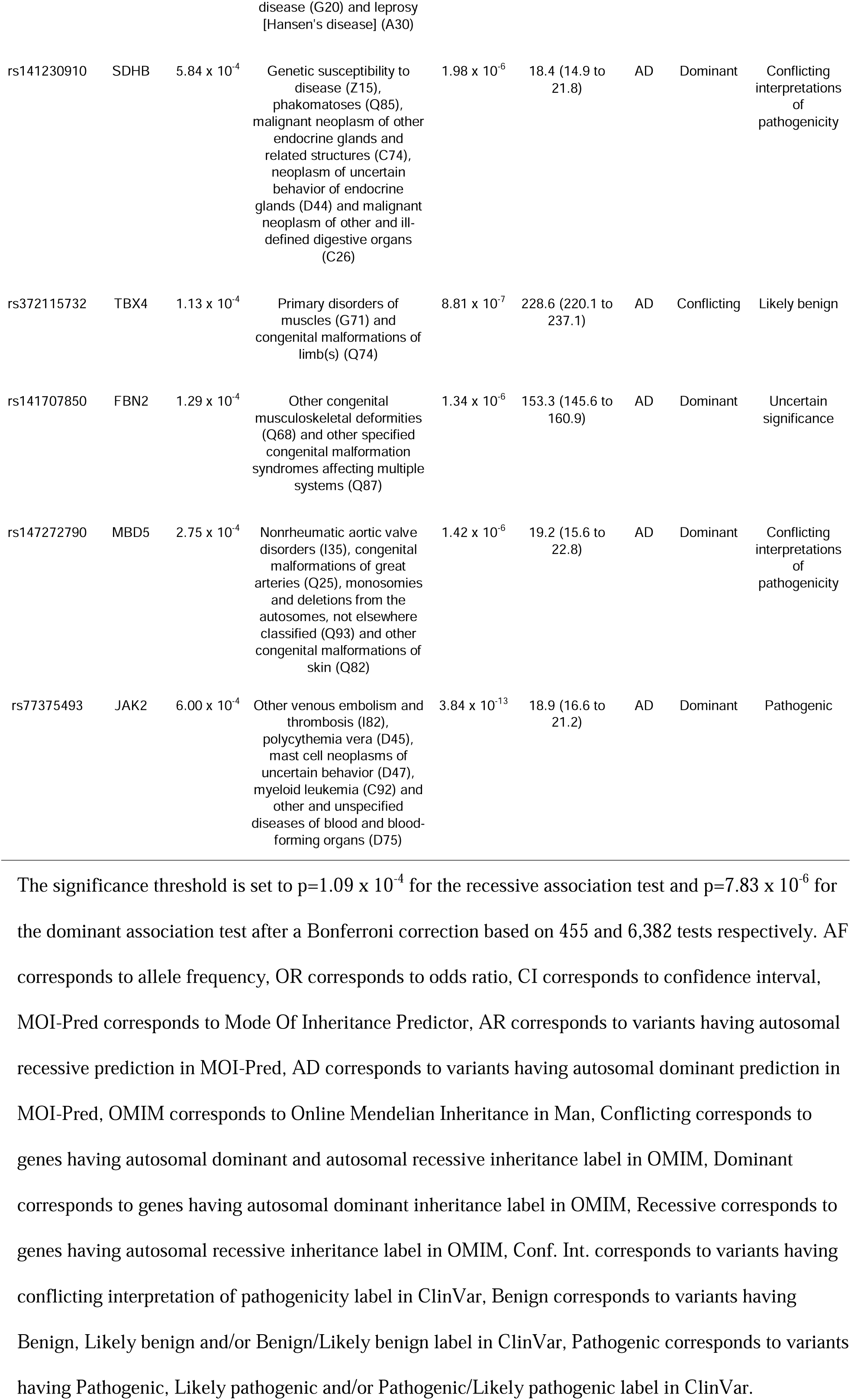
Description of significant variant associations with disease.

## Discussion

Here we present MOI-Pred, a computational tool that jointly predicts pathogenicity and mode of inheritance for missense variants. Our tool uses a random forest classifier trained on known variants to combine multiple sources of annotation into a single prediction. Compared to other existing methods, MOI-Pred benefits from several key innovations. First and foremost, where most methods produce a binary prediction of pathogenic or benign, our method produces a three-way prediction, classifying each variant as pathogenic for AR disease, pathogenic for AD disease, or benign. In particular, while many existing methods perform well at predicting pathogenic variants in AD disease (e.g. O/E ^41^, CADD ^44^, phyloP ^45^, etc.), MOI-Pred specifically targets the problem of discriminating AR pathogenic variants from benign, a long-lasting issue in genetics unattended by current annotation pipelines. Only one pre-existing method makes three-class predictions, MAPPIN^38^. MOI-Pred performs substantially better than MAPPIN at identifying variants associated with recessive-acting diseases in both prediction performance (Precision=0.79 on the training set for MAPPIN recessive predictions based on Gosalia et al. 2017^38^) and when testing on real-world clinical data in the current study.

Second, MOI-Pred combines evolutionary and functional annotations on both the gene and variant level to produce a combined variant-level prediction. This gives an important advantage in predicting mode of inheritance, since different annotation sources are known to have different error profiles. In particular, it has recently been shown that evolutionary scores are primarily sensitive to heterozygote effects, making these methods very likely to misclassify AR pathogenic variants as benign^34,35^. Most predictors of pathogenicity rely primarily on these scores, and therefore may be systematically insensitive to AR pathogenic variants. By combining multiple different scores in a random forest framework, MOI-Pred is able to learn which scores are most sensitive to each mode of inheritance. For example, O/E, which relies exclusively on evolutionary constraint, is very likely to confuse AR with benign, while MutPred, which incorporates biophysical properties of proteins^46,47^, is more likely to categorize AR variants as pathogenic. Accordingly, in our method, O/E is an important feature separating AD from benign, while MutPred is an important feature separating AR from benign.

Third, MOI-Pred is trained with a population-derived list of benign variants, and validated on novel population-derived benign variants unknown to its constituent scores. Because the most difficult classification task is distinguishing AR from benign, the choice of benign training data is vitally important. Since we use clinically validated pathogenic variants for training, it is tempting to use clinically validated benign variants as well, but this can bias the training set. These were suspected being pathogenic at some point and therefore may have features that are not typical of benign variants^48^. The ideal source of benign variants should be found at sufficiently high frequency in a healthy human population^13,49^. We used frequency-matched variants from a large population database (gnomAD) as presumed benign controls in our training set, an approach that has been used by previous methods such as CADD^44^ and VEST3^50,51^. However, using these variants introduces an additional problem: population genetics scores used as components in our prediction model are often themselves derived from the same populations, introducing bias and the risk of overfitting^52^. We addressed this problem by using common variants from a recently published cohort of healthy Japanese adults as a validation set. At the time of analysis, this population had not yet been incorporated into widely used population databases, and all genetics scores were therefore naïve to it. Our classifier performed better on these population-derived benign variants than an equivalent classifier trained on clinically validated benign variants, and also performed well on clinically validated benign variants.

Finally, we validated our method by using it to predict disease case-control status in EHR data from the Bio*Me* biobank^43^. We demonstrated that our predictions of mode of inheritance are significantly associated with the likelihood of carriers developing Mendelian disease in a real-world clinical setting. This is true for variants annotated as pathogenic, variants of unknown significance, as well as novel and ancestry-specific variants. This analysis also revealed that some variants with a Benign prediction appear to protect against disease, particularly in variants with “Uncertain significance” (Figure 4C). Since the “Uncertain significance” ClinVar class necessarily contains variants without clear evidence for or against pathogenicity, it is possible that disease modifier variants or variants with protective effects in heterozygous or homozygous form (underdominance or overdominance) may frequently be classified as “Uncertain significance.” Similarly, these variants lack normal signatures of natural selection and so may be likely to receive a Benign prediction in MOI-Pred. In general, the properties of these variants are not well understood, including whether they can be predicted by computational methods, and further investigation is warranted^53^. We also found individual variants where the prediction made by MOI-Pred differed from their clinical annotations using ClinVar/OMIM, and we verified using the same EHR database that both the pathogenicity and the mode of inheritance predicted by our method is likely to be correct. These analyses demonstrate the applicability of our method to real clinical data and decision-making particularly with respect to large-scale electronic health systems. Such clinical validation is only possible thanks to increasingly available EHR-linked biobanks, and we anticipate it being applied more broadly to variant prediction tools in the future.

Our method has several limitations and areas for future work. First, it remains uncertain whether the performance we observe in the test and validation sets will hold in real applications. Many existing tools have reported similarly high performance in their authors’ internal testing and lower performance in unbiased replication analyses^12^. Many have also failed to find clinical utility despite numerically high performance^54,55^. Our EHR-based clinical validation suggests that results will hold^56^ in real-world clinical data, but validation in other clinical datasets and by other groups is needed. Second, our three-way predictions, though more complete than typical binary predictions, do not completely account for all forms of mode of inheritance. Phenomena such as incomplete dominance, overdominance, and heterozygote advantage, all of which are well documented in human disease^56-58^, are unaccounted for in our simple recessive-dominant-benign classification. Likewise, mode of inheritance itself is far from the only refinement that can be added to variant effect predictions. The field would benefit enormously from methods to predict gain-of-function variants, disease suppressor variants, or uniparental imprinted variants, to name just a few. Third, there is room for innovation and improvement in the method. We used only a subset of applicable ML methods and available features. Furthermore, we focus only on missense variants, primarily because this is the largest class of variation for clinical variants and most applicable to mode of inheritance prediction. Most coding annotations are available for missense variants, while a much smaller number are available for other forms of variation including synonymous, loss-of-function, non-coding, or multi-nucleotide variants. Ultimately, MOI-Pred is meant to be used in combination with other methods to form a holistic picture of the effects of variants. This is true for even the most widely-used prediction methods, which are rarely relied on individually. We believe that this method, together with MAPPIN and others, will enable variant function prediction to go beyond a binary prediction of pathogenicity so that the picture of variant effects formed by computational annotation begins to resemble the true complexity of actual phenotypes.

## Methods

### Variant collection

Missense variants from publicly available resources were used to generate all datasets. For the training set, pathogenic variants were obtained from ExoVar^16^. Presumed non-pathogenic variants were selected from the Genome Aggregation Database (gnomAD)^41^ v2.1.1. GnomAD variants were chosen to match the allele frequency (AF) of pathogenic variants to within 0.1%, based on minor allele frequency in the entire gnomAD population; singletons were chosen to match variants not present in gnomAD. For the validation set, pathogenic variants with review status “reviewed by expert panel” were selected from ClinVar^27^ (release June 2020), and presumed non-pathogenic variants were selected from GEM^42^, defined as variants with AF >= 1% in GEM and absent or singleton in gnomAD. Gene-level mode of inheritance information for pathogenic variants was obtained from OMIM^26^ (release May 2020).

### Variant annotation

Variants were characterized using functional and evolutionary information. ANNOVAR^59^ was used to annotate variant-level features. We included all available features from ANNOVAR that could be applied to all or nearly all missense variants. This includes 15 features built on evolutionary information (e.g. phyloP^45^, FATHMM^60^, GERP^61^, PROVEAN^62^, etc.), 42 features built on both evolutionary and functional information (e.g. M-CAP^63^, CADD^44^, VEST3^50^, MutationTaster^64^, etc.) and 14 population frequency features (e.g. cg69^65^, Kaviar^66^, GME^67^, etc.). We added to this, 7 gene-level features which were retrieved manually from their original sources. This includes 2 gene level features built on evolutionary information (OE score^41^ and s_het^68^), 4 gene level features built on functional information (Episcore^69^, AD rank^70^, StringAD and StringAR^71^) and 1 gene level feature combining the two annotations (HI^72^). The full list of features and their source of information can be found in **Supplementary Table 1**.

### Data trimming and imputation

Features with more than 60% missing values and/or high correlation (Pearson’s r >= 0.8) in the training set were removed. In two correlated features, the one with higher mean absolute correlation across all features was removed. Variants with more than 60% missing values in the remaining set of features were removed from both the training and external validation sets. Missing values were imputed first on variant-level features. The resulting dataset was then used to impute gene-level features, ensuring low intra-gene variation in these annotations. A random forest-based algorithm (missForest v1.4^73^) was used for both imputations. The final dataset was comprised of 30 features on 5,872 and 1,526 variants from the training and external validation sets respectively.

### Workflow to train inheritance prediction models

A machine learning (ML) approach was used to develop mode of inheritance prediction models. To minimize sampling biases, 100 models were trained, tested and validated using random sets of variants. The workflow is described below for a single iteration. A random sample of 90% of available Dominant variants from the training set plus equal numbers of Recessive and Benign variants constituted a balanced Train set. The remaining variants from the training set were used to sample a balanced 10% Test set. The Validation set consisted of all Dominant variants available in the external validation dataset plus equal numbers of randomly sampled Recessive and Benign variants. Scaling and feature selection (using a wrapper random forest-based approach, recursive feature elimination) were performed on the Train set using the caret package v6.0.84^74^ available in R, then applied accordingly to the Test and Validation sets. A three-class (Recessive, Dominant, Benign) random forest algorithm^75^ was then fitted to the Train set using 10-fold cross validation to optimize parameter tuning and limit overfitting. Three two-class random forest algorithms (Dominant vs. Recessive, Dominant vs. Benign, Recessive vs. Benign) were fitted in parallel for subsequent feature importance analyses. Mode of inheritance label was then predicted on the Test and Validation sets to compute performance metrics. This entire procedure was repeated 100 times; reported performance statistics (see Results) correspond to the mean and standard deviation (SD) across all 100 runs.

The AUROC was calculated using the pROC package v1.14.0^76^ available in R v3.5.3^77^. To obtain a per-class discrimination metric the remaining two labels were treated as negative classes. Accuracy, sensitivity, specificity and positive/negative predictive values (PPV/NPV) as well as the ML framework was implemented using the caret package.

Three-way variant-level mode of inheritance predictions (pathogenic for autosomal recessive (AR) disease, pathogenic for autosomal dominant (AD) disease, or benign) for all possible missense variants in the human genome build hg38 are available at https://github.com/rondolab/MOI-Pred/.

### Clinical validation of inheritance prediction models in electronic health records

A single three-class random forest algorithm was fitted, tested and validated as described above to predict mode of inheritance in genotype data from 29,981 individuals in the Bio*Me* biobank^43^. Bio*Me* is a multiethnic, EHR-linked, clinical care biobank of more than 60,000 samples from individuals recruited at the Mount Sinai Health System between 2007 and 2015. Participants were genotyped using the Illumina Global Screening Array, imputation was performed using the 1000 Genomes Phase 3 reference panel, and genetic ancestry was determined through k-means clustering of principal components. Longitudinal biomedical traits including diagnostic codes and laboratory test results were obtained mainly through ambulatory care practices resulting in a high median number of encounters per patient^78^. Only variants present in ClinVar (release June 2020) were considered for posterior analyses. ClinVar’s phenotype information was mapped to 456 categories of International Classification of Disease 10 (ICD-10) diagnostic codes using information from the Systematized Nomenclature of Medicine Clinical Terms (SNOMED-CT)^79^ and Orphanet^80^.

Contingency table analyses were performed to test recessive, dominant and benign models on variants predicted with the corresponding mode of inheritance. Each table evaluates a subset of variants having the same inheritance prediction, same clinical significance label in ClinVar, and mapped to the same set of billing codes. An individual was considered “affected” if diagnosed with an ICD-10 code mapped to the above-mentioned subset of variants. Likewise, an individual was considered a “carrier” if homozygous for the pathogenic allele for the recessive model, homozygous for the pathogenic allele or heterozygous for the dominant model, and homozygous for the pathogenic allele or heterozygous for the benign model. An individual was considered a ‘non-carrier’ if heterozygous for the recessive model, homozygous for the non-pathogenic allele for the dominant model, and homozygous for the non-pathogenic allele for the benign model. Each 2×2 table of carrier status vs. phenotype case/control status was restricted to individuals from independent ancestries and weighted by the prevalence of the corresponding set of ICD-10 codes in each of the ancestries in the Bio*Me* EHR data. The analysis was repeated twice, once for variants with predicted mode of inheritance corresponding to the model (e.g. variants predicted Recessive when evaluating on the recessive model) and once for all variants not predicted in the respective model (e.g. variants predicted Benign and Dominant when evaluating on the recessive model). Furthermore, a secondary analysis restricting to variants with ClinVar review status of two stars or higher was performed.

A CMH test was applied to the ancestry-specific tables from the same predicted mode of inheritance and clinical significance to obtain OR, 95% CI and corresponding p-values. The results were then aggregated across ancestries using an inverse variance meta-analysis. A Q-test was performed to evaluate heterogeneity between ORs of variants predicted and not predicted in the respective models. The stats package v3.6.2^77^ was used to perform the CMH test and the metafor package v3.0.2^81^ was used for the Q-test.

### Single nucleotide variant association discovery

A total of 433 groups of ICD-10 codes were tested for dominant and recessive association with 6,382 variants present in ClinVar, having an ICD-10 code mapping and MOI-Pred prediction. The analysis was performed in individual ancestries (European-American, African-American, Hispanic-American and other ancestries) and meta-analyzed using plink v1.9^82^. Whole exome sequencing data and EHR from the Bio*Me* biobank were used in the analysis; 10 principal components were used as covariates to account for population stratification.

## Supporting information

Supplemental methods and results.

## Data Availability

All data produced are available at https://github.com/rondolab/MOI-Pred

## Author Contributions

Dr. Do, Dr. Jordan and Mr. Petrazzini had full access to all of the data in the study and take responsibility for the integrity of the data and accuracy of the data analysis.

*Concept and design:* Petrazzini, Jordan, Do.

*Acquisition, analysis, or interpretation of the data:* All authors.

*Drafting of the manuscript:* Petrazzini, Balick, Forrest, Rocheleau, Jordan and Do.

*Critical revision of the manuscript for important intellectual concept:* All authors.

*Statistical analysis:* Petrazzini, Rocheleau, Jordan, Do.

*Obtained funding:* Do.

*Administrative, technical, or material support:* Cho, Do.

*Supervision:* Jordan and Do.

## Disclaimer

The content is solely the responsibility of the authors and does not necessarily represent the official views of the National Institutes of Health.

## Data sharing statement

Not applicable.

