## Supplemental methods and results. for "Prediction of recessive inheritance for missense variants in human disease"

### Supplementary Material

#### Figures

**Supplementary Figure 1.** Detailed study design and machine learning workflow.

**Supplementary Figure 2.** Bar-plots showing sensitivity and specificity for 3-class mode of inheritance prediction models per ClinVar review status.

**Supplementary Figure 3.** Bar-plots showing sensitivity and specificity for 3-class mode of inheritance prediction models per ClinVar review status on European American-specific variants.

**Supplementary Figure 4.** Bar-plots showing sensitivity and specificity for 3-class mode of inheritance prediction models per ClinVar review status on African American-specific variants.

**Supplementary Figure 5.** Bar-plots showing sensitivity and specificity for 3-class mode of inheritance prediction models per ClinVar review status on Hispanic American-specific variants.

**Supplementary Figure 6.** Bar-plot showing feature importance on 2-class (Benign - Dominant) mode of inheritance prediction models.

**Supplementary Figure 7.** Bar-plot showing feature importance on 2-class (Benign - Recessive) mode of inheritance prediction models.

**Supplementary Figure 8.** Bar-plot showing feature importance on 2-class (Dominant - Recessive) mode of inheritance prediction models.

**Supplementary Figure 9.** Forest plots showing P values and effect sizes for association with disease in variants predicted Recessive (A), Dominant (B) or Benign (C) and having 2-star review statis or higher in ClinVar.

**Supplementary Figure 10.** Forest plots showing P values and effect sizes for association with disease in variants predicted Recessive (A), Dominant (B) or Benign (C) and having 2-star review statis or higher in ClinVar.

#### Tables

**Supplementary Table 1.** Complete list of features tested in the machine learning workflow and the corresponding information used to generate each feature.

**Supplementary Table 2.** Counts of variants missing mode of inheritance information in ClinVar.

**Supplementary Table 3.** Counts of variants used in the clinical validation on the Bio*Me* biobank.

**Supplementary Table 4.** Performance metrics for the 3-class mode of inheritance prediction model used in the clinical validation on the Bio*Me* biobank.

**Supplementary Table 5.** Detailed P values and effect sizes for the disease association test on variants predicted Recessive in the Bio*Me* biobank.

**Supplementary Table 6.** Detailed P values and effect sizes for the disease association test on variants predicted Dominant in the Bio*Me* biobank.

**Supplementary Table 7.** Detailed P values and effect sizes for the disease association test on variants predicted Benign in the Bio*Me* biobank.

**Supplementary Table 8.** Detailed P values and effect sizes for the disease association test on variants predicted Recessive in the Bio*Me* biobank, restricted to 2-star review status or higher.

**Supplementary Table 9.** Detailed P values and effect sizes for the disease association test on variants predicted Dominant in the Bio*Me* biobank, restricted to 2-star review status or higher.

**Supplementary Table 10.** Detailed P values and effect sizes for the disease association test on variants predicted Benign in the Bio*Me* biobank, restricted to 2-star review status or higher.

**Supplementary Table 11.** Detailed P values and effect sizes for the disease association test on variants predicted Recessive by MAPPIN in the Bio*Me* biobank.

**Supplementary Table 12.** Detailed P values and effect sizes for the disease association test on variants predicted Dominant by MAPPIN in the Bio*Me* biobank.

**Supplementary Table 13.** Detailed P values and effect sizes for the disease association test on variants predicted Benign by MAPPIN in the Bio*Me* biobank.

**Supplementary Table 14.** Description of significant SNP associations with disease on African Americans.

**Supplementary Table 15.** Description of significant SNP associations with disease on European Americans.

**Supplementary Table 16.** Description of significant SNP associations with disease on Hispanic Americans.

**Supplementary Table 17.** Description of significant SNP associations with disease on other ancestries.

**Supplementary Figure 1**: Detailed study design and machine learning workflow.

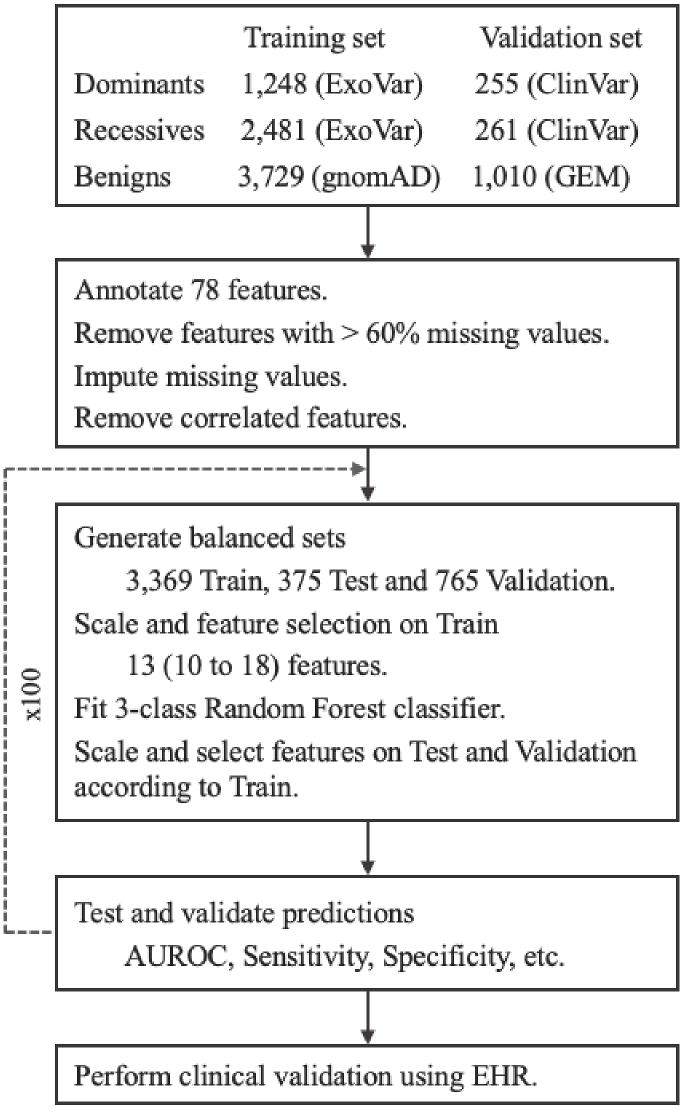


Train and Test sets correspond to the 90% and 10% balanced datasets, built from ExoVar and gnomAD variants, used for training and testing respectively. Validation set corresponds to the balanced dataset, built from ClinVar and GEM variants, used for external validation. Number of features is reported as median (min to max) across 100 models. AUROC corresponds to area under the receiver operator characteristic curve. EHR corresponds to electronic health records.

**Supplementary Figure 2**: Bar-plots showing sensitivity and specificity for 3-class mode of inheritance prediction models per ClinVar review status.

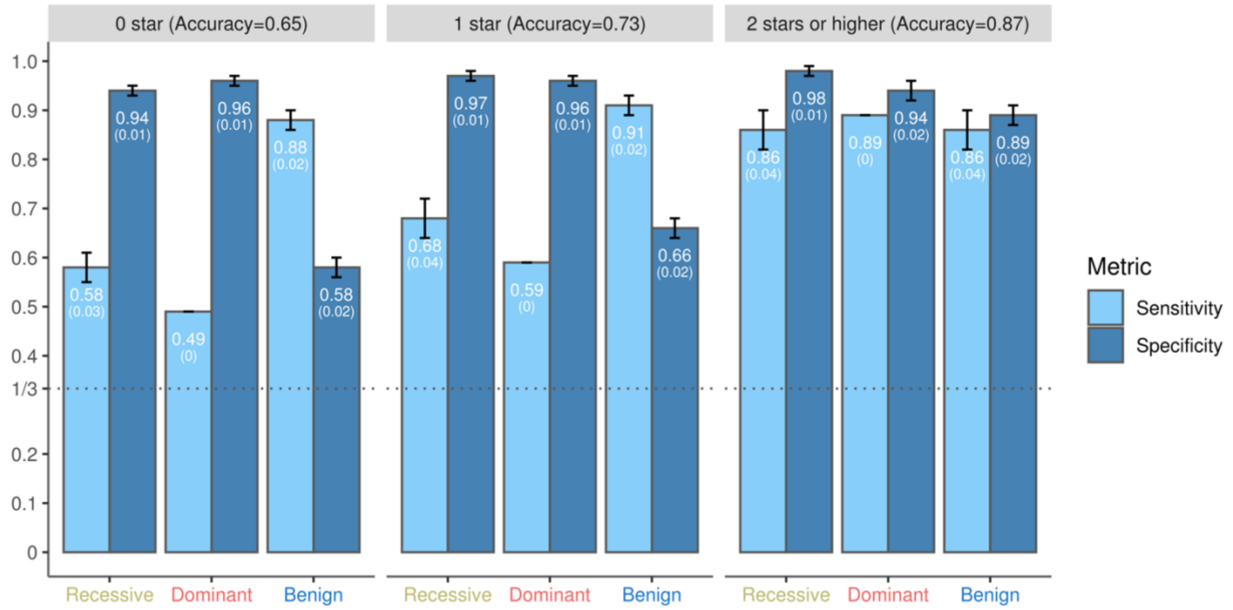


0 Star review status in ClinVar includes “no assertion for the individual variant”, “no assertion criteria provided” and “no assertion provided”. 1 Star includes “criteria provided, single submitter” and “criteria provided, conflicting interpretations”. 2 Stars or higher includes “criteria provided, multiple submitters, no conflicts”, “reviewed by expert panel” and “practice guideline”.

**Supplementary Figure 3**: Bar-plots showing sensitivity and specificity for 3-class mode of inheritance prediction models per ClinVar review status on European American-specific variants.

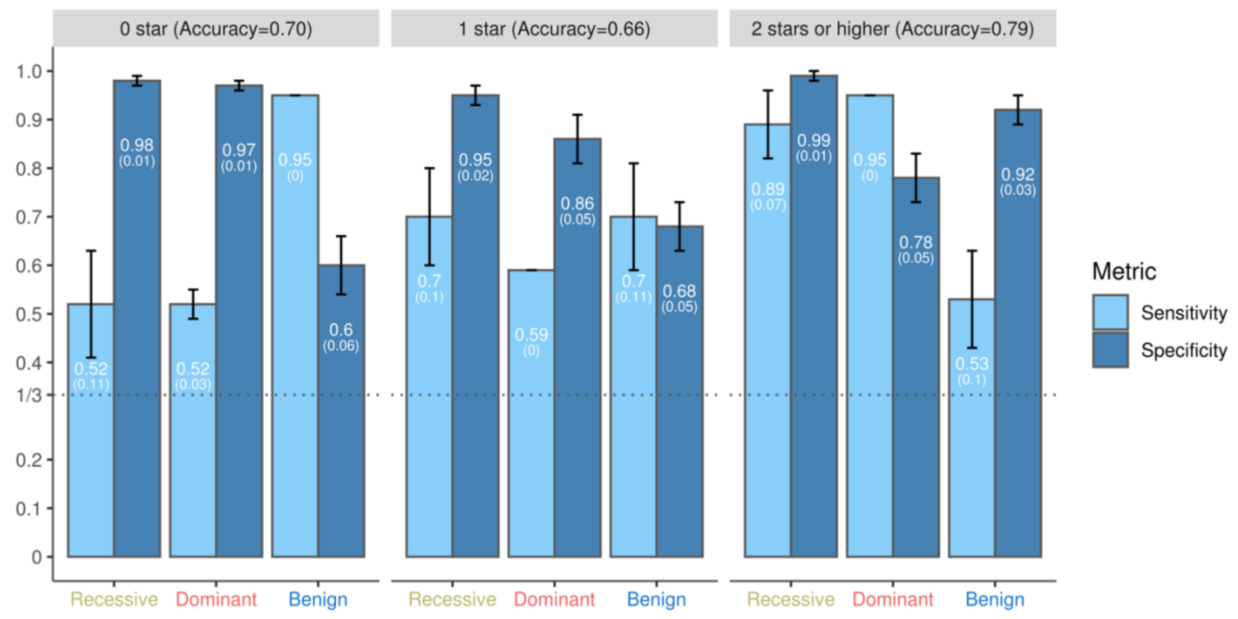


Patients from European American ancestry were defined based on questionnaire data. 0 Star review status in ClinVar includes “no assertion for the individual variant”, “no assertion criteria provided” and “no assertion provided”. 1 Star includes “criteria provided, single submitter” and “criteria provided, conflicting interpretations”. 2 Stars or higher includes “criteria provided, multiple submitters, no conflicts”, “reviewed by expert panel” and “practice guideline”.

**Supplementary Figure 4**: Bar-plots showing sensitivity and specificity for 3-class mode of inheritance prediction models per ClinVar review status on African American-specific variants.

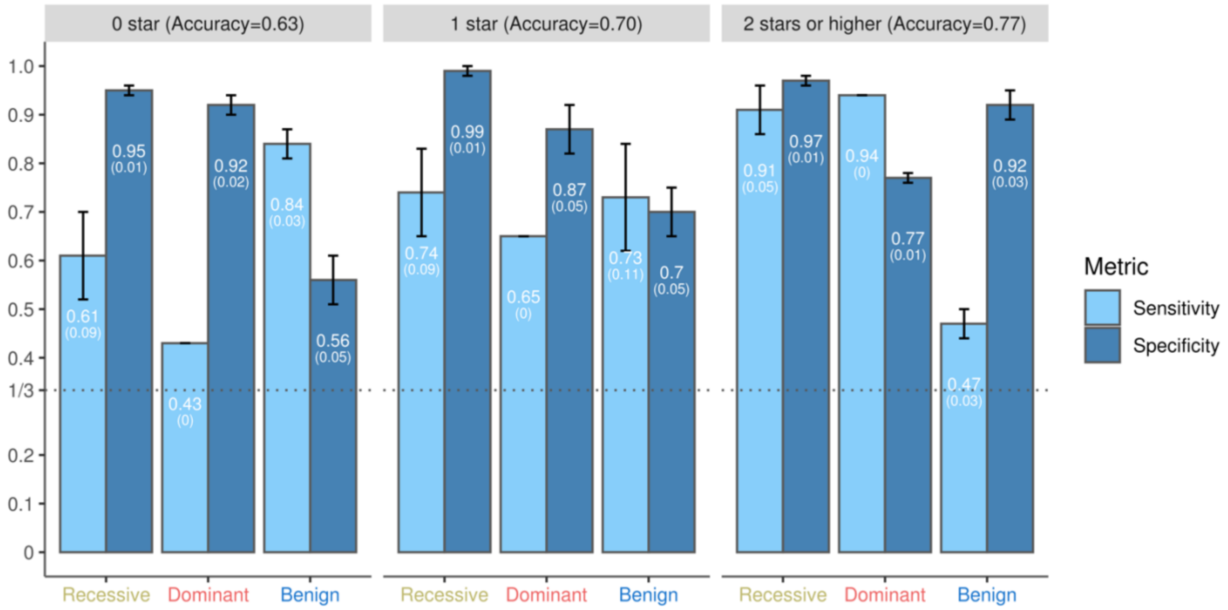


Patients from African American ancestry were defined based on questionnaire data. 0 Star review status in ClinVar includes “no assertion for the individual variant”, “no assertion criteria provided” and “no assertion provided”. 1 Star includes “criteria provided, single submitter” and “criteria provided, conflicting interpretations”. 2 Stars or higher includes “criteria provided, multiple submitters, no conflicts”, “reviewed by expert panel” and “practice guideline”.

**Supplementary Figure 5**: Bar-plots showing sensitivity and specificity for 3-class mode of inheritance prediction models per ClinVar review status on Hispanic American-specific variants.

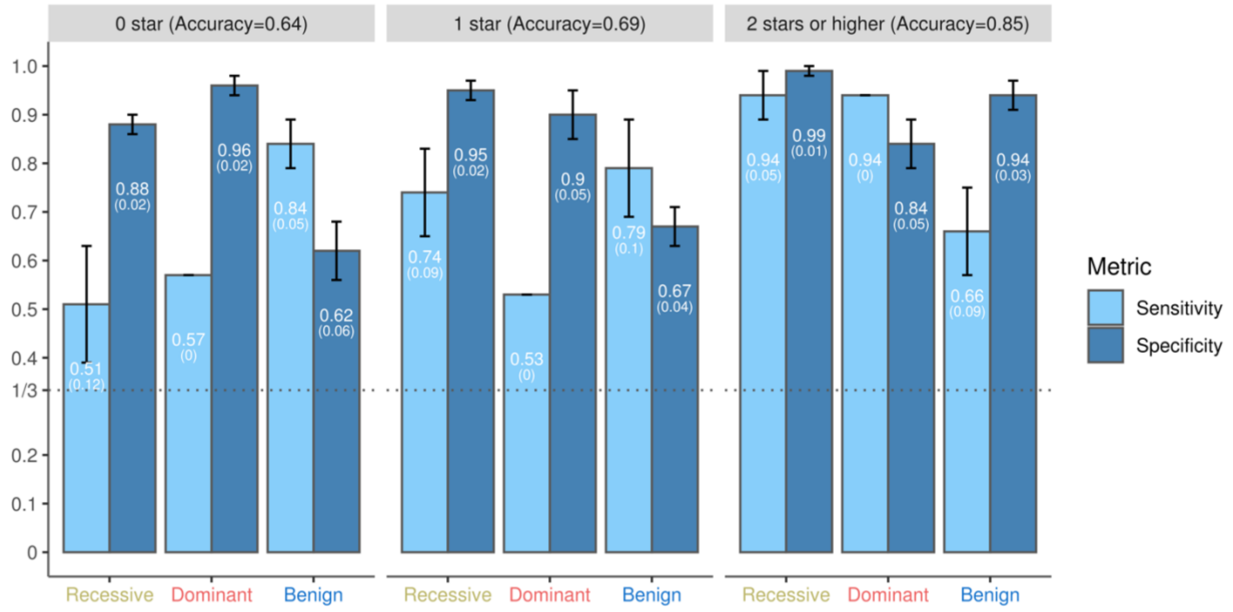


Patients from Hispanic American ancestry were defined based on questionnaire data. 0 Star review status in ClinVar includes “no assertion for the individual variant”, “no assertion criteria provided” and “no assertion provided”. 1 Star includes “criteria provided, single submitter” and “criteria provided, conflicting interpretations”. 2 Stars or higher includes “criteria provided, multiple submitters, no conflicts”, “reviewed by expert panel” and “practice guideline”.

**Supplementary Figure 6**: Bar-plot showing feature importance on 2-class (Benign - Dominant) mode of inheritance prediction models.

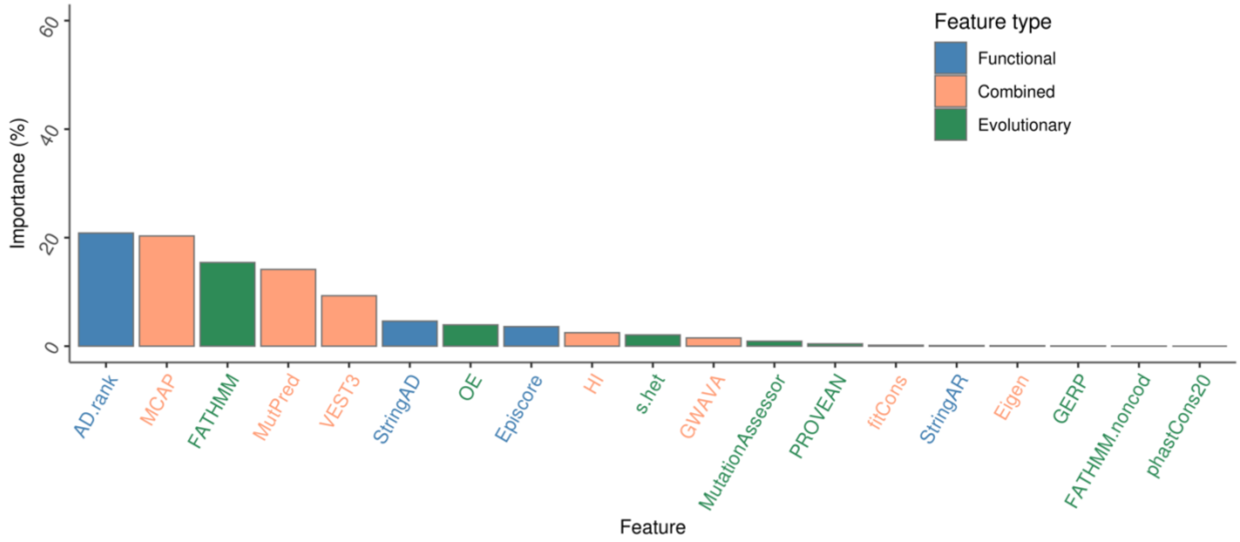


Feature importance is reported as the median across 100 models.

**Supplementary Figure 7**: Bar-plot showing feature importance on 2-class (Benign - Recessive) mode of inheritance prediction models.

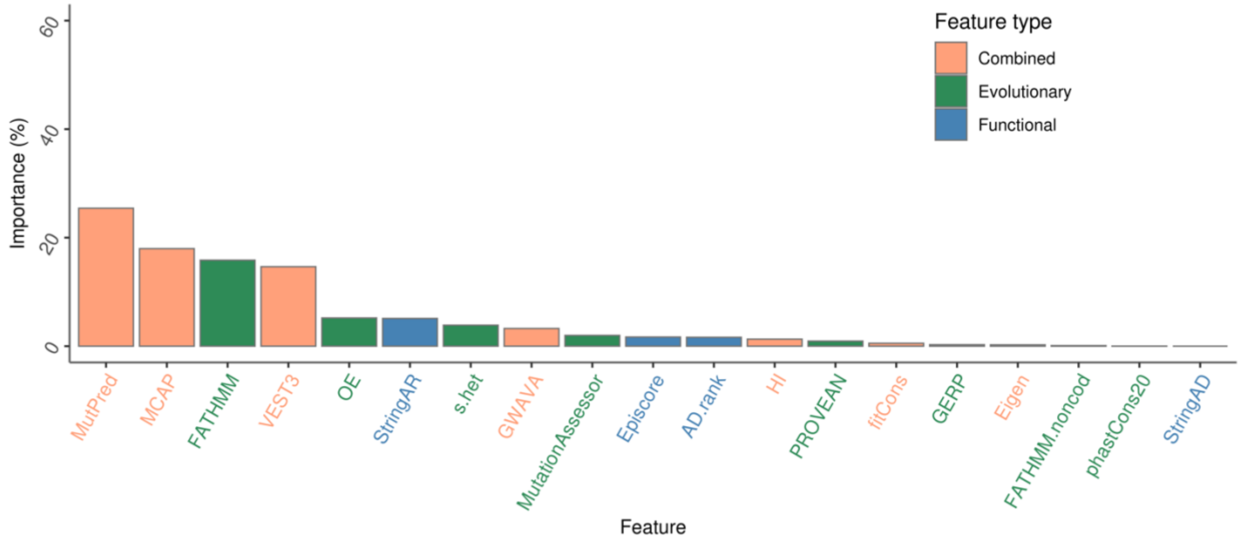


Feature importance is reported as the median across 100 models.

**Supplementary Figure 8**: Bar-plot showing feature importance on 2-class (Dominant - Recessive) mode of inheritance prediction models.

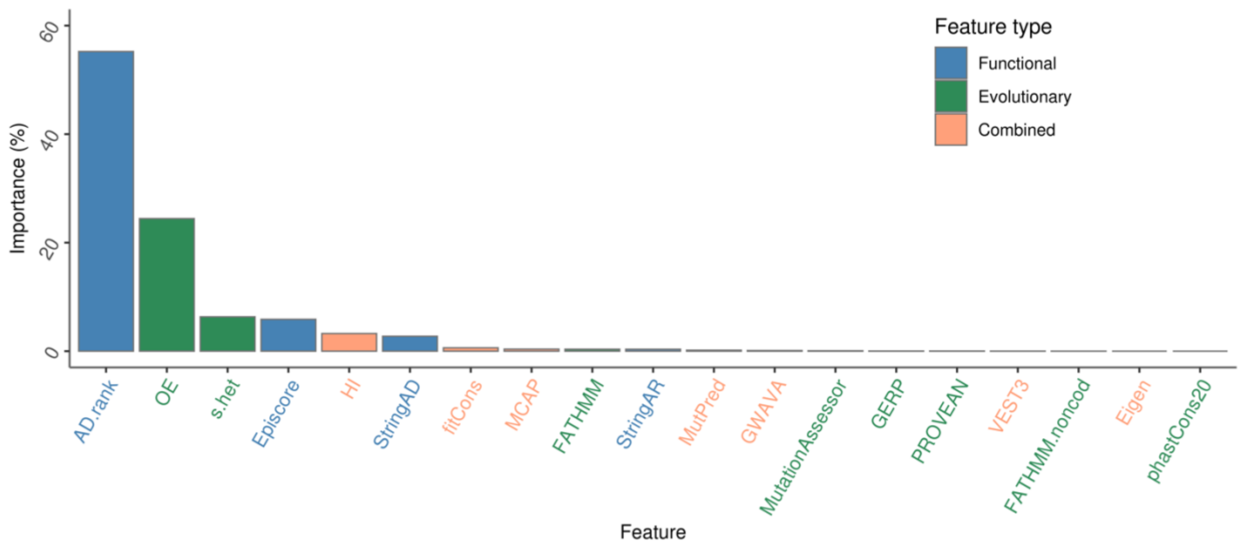


Feature importance is reported as the median across 100 models.

**Supplementary Figure 9**: Forest plots showing P values and effect sizes for association with disease in variants predicted Recessive (A), Dominant (B) or Benign (C) and having 2-star review statis or higher in ClinVar.

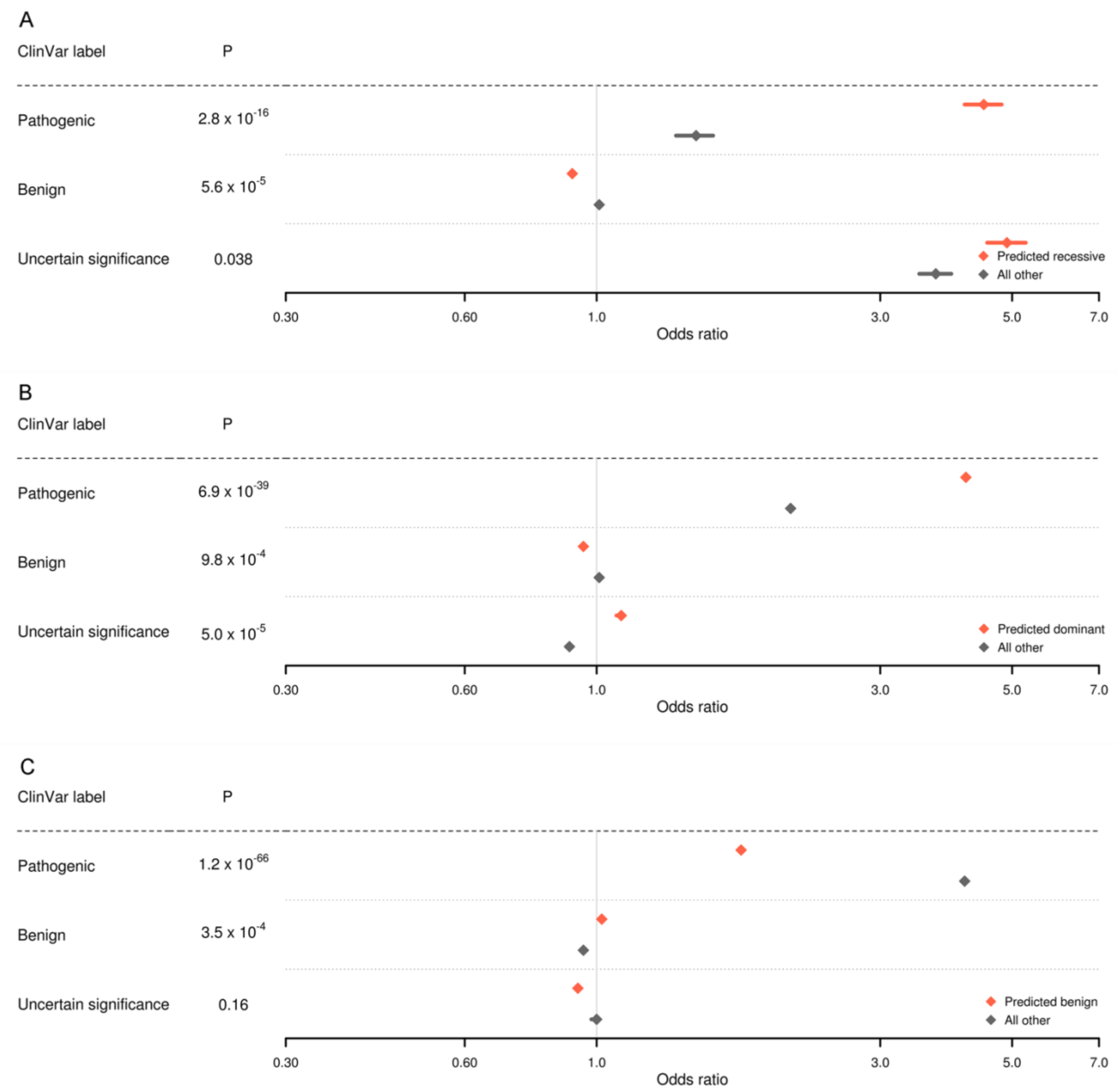

Effect sizes (Odds ratios) and 95% confidence intervals were obtained for individual ancestries using a Cochran-Mantel-Haenszel (CMH) test. The reported effect sizes correspond to an inverse variance meta-analysis across ancestries. P values for heterogeneity between Odds ratios are derived from a Q-test Test set.

**Supplementary Figure 10**: Forest plots showing P values and effect sizes for association with disease in variants predicted Recessive (A), Dominant (B) or Benign (C) by MAPPIN.

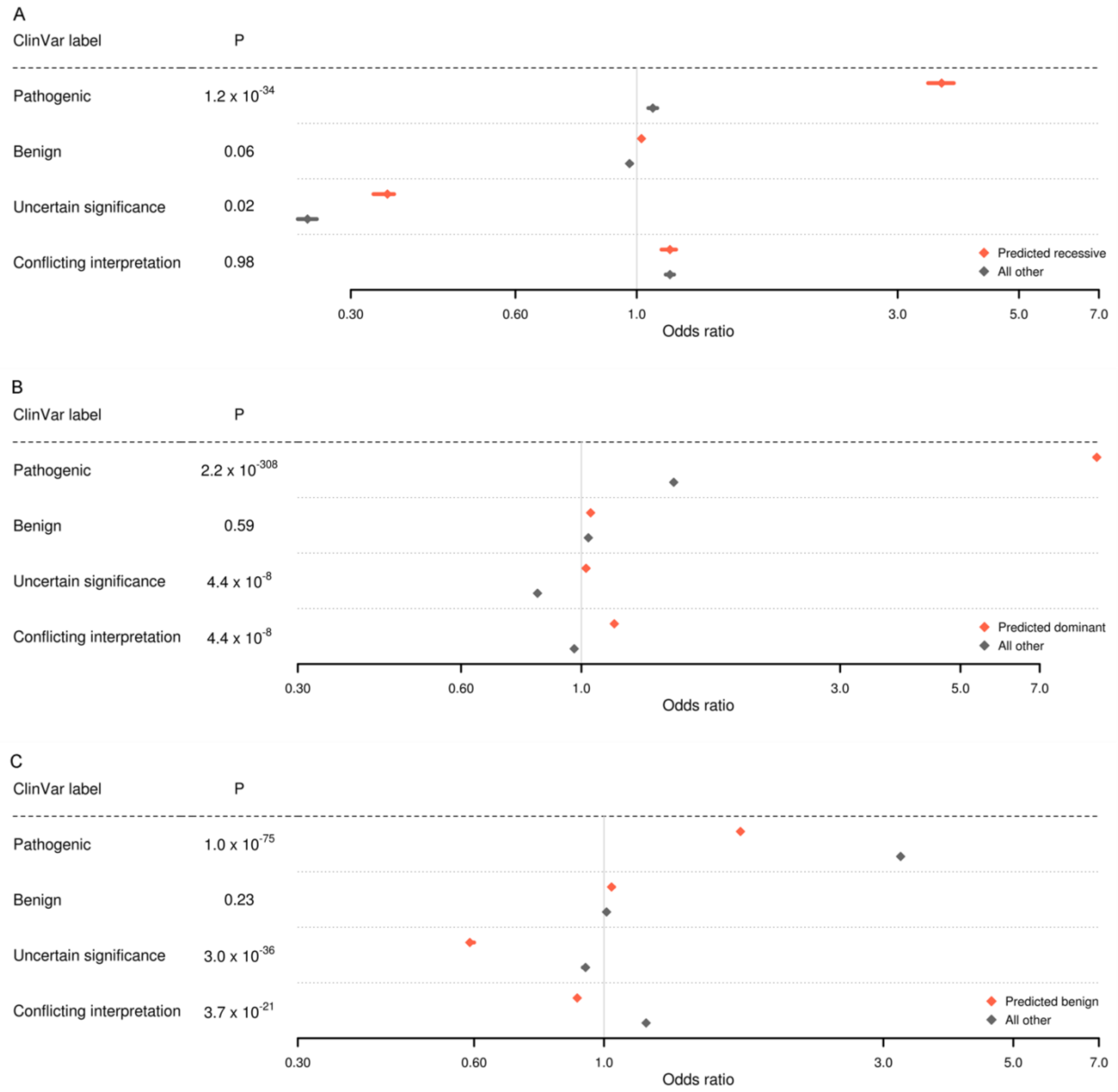

Effect sizes (Odds ratios) and 95% confidence intervals were obtained for individual ancestries using a Cochran-Mantel-Haenszel (CMH) test. The reported effect sizes correspond to an inverse variance meta-analysis across ancestries. P values for heterogeneity between Odds ratios are derived from a Q-test Test set.

**Supplementary Table 1.** Complete list of features tested in the machine learning workflow and the corresponding information used to generate each feature.

| Feature | Information used |
|---|---|
| abroam | Allele frequency |
| cg60 | Allele frequency |
| cg46 | Allele frequency |
| SIFT | Evolutionary |
| LRT | Evolutionary |
| MutationTaster | Combined |
| MutationAssessor | Evolutionary |
| FATHMM | Evolutionary |
| PROVEAN | Evolutionary |
| MetaSVM | Combined |
| MetaLR | Combined |
| MCAP | Combined |
| MutPred | Combined |
| fathmm | Evolutionary |
| Eigen | Combined |
| GenoCanyon | Combined |
| fitCons | Combined |
| GERP | Evolutionary |
| phyloP100 | Evolutionary |
| phyloP46 | Evolutionary |
| phyloP20 | Evolutionary |
| phastCons100 | Evolutionary |
| phastCons20 | Evolutionary |
| SiPhy29 | Evolutionary |
| GME | Allele frequency |
| GME NWA | Allele frequency |
| GME NEA | Allele frequency |
| GME AP | Allele frequency |

| | |
|---|---|
| GME Israel | Allele frequency |
| GME SD | Allele frequency |
| GME TP | Allele frequency |
| GME CA | Allele frequency |
| HRC | Allele frequency |
| HRC non 1000G | Allele frequency |
| Kaviar | Allele frequency |
| GWAVA | Combined |
| PVS1 | Combined |
| PS1 | Combined |
| PS2 | Combined |
| PS3 | Combined |
| PS4 | Combined |
| PM1 | Combined |
| PM2 | Combined |
| PM3 | Combined |
| PM4 | Combined |
| PM5 | Combined |
| PM6 | Combined |
| PP1 | Combined |
| PP2 | Combined |
| PP3 | Combined |
| PP4 | Combined |
| PP5 | Combined |
| BA1 | Combined |
| BS1 | Combined |
| BS2 | Combined |
| BS3 | Combined |
| BS4 | Combined |
| BP1 | Combined |
| BP2 | Combined |
| BP3 | Combined |
| BP4 | Combined |

| | |
|---|---|
| BP5 | Combined |
| BP6 | Combined |
| BP7 | Combined |
| Polyphen2 HDIV | Combined |
| Polyphen2 DVAR | Combined |
| VEST3 | Combined |
| CADD | Combined |
| REVEL | Combined |
| OE | Evolutionary |
| s_het | Evolutionary |
| Episcore | Functional |
| StringAR | Functional |
| StringAD | Functional |
| HI | Combined |
| AD_rank | Functional |

The feature space used includes 71 variant-level features and 7 gene-level features. Variant-level features are divided in 15 features built using evolutionary information, 42 features built using both evolutionary and functional information and 14 population frequency features. Gene-level features are divided in 2 features built using evolutionary information, 4 features built using functional information and 1 feature combining the two annotations.

**Supplementary Table 2.** Counts of variants missing mode of inheritance information in ClinVar.

| ClinVar label | Total | Missing MOI information (#) | Missing MOI information (%) |
|---|---|---|---|
| Pathogenic | 140,090 | 49,745 | 35.51 |
| Benign | 288,873 | 119,532 | 41.38 |
| Uncertain significance | 347,328 | 122,600 | 35.30 |
| Conflicting interpretation | 41,693 | 15,923 | 38.19 |
| All | 817,984 | 307,800 | 37.63 |

Counts correspond to ClinVar release June 2020. MOI corresponds to mode of inheritance.

**Supplementary Table 3.** Counts of variants used in the clinical validation on the Bio*Me* biobank.

| ClinVar label | All missense | 2-star or higher |
|---|---|---|
| Pathogenic | 2,301 | 1,047 |
| Benign | 9,865 | 6,303 |
| Uncertain significance | 35,629 | 11,784 |
| Conflicting interpretation | 8,911 | NA |
| All | 56,706 | 19,134 |

Variants obtained from Bio*Me* exome data were filtered if missing >60% annotations, not present in ClinVar and/or missing ICD-code mapping.

**Supplementary Table 4.** Performance metrics for the 3-class mode of inheritance prediction model used in the clinical validation on the Bio*Me* biobank.

| Dataset | Accuracy | Recessive sensitivity | Dominant sensitivity | Benign sensitivity | Recessive specificity | Dominant specificity | Benign specificity |
|---|---|---|---|---|---|---|---|
| Train | 0.93 | 0.94 | 0.96 | 0.90 | 0.96 | 0.97 | 0.96 |
| Test | 0.82 | 0.76 | 0.78 | 0.92 | 0.94 | 0.98 | 0.81 |
| Validation | 0.88 | 0.85 | 0.85 | 0.95 | 0.99 | 0.99 | 0.85 |

Train and Test sets correspond to the 90% and 10% balanced datasets, built from ExoVar and gnomAD variants, used for training and testing respectively. Validation set corresponds to the balanced dataset, built from ClinVar and GEM variants, used for external validation.

**Supplementary Table 5.** Detailed P values and effect sizes for the disease association test on variants predicted Recessive in the Bio*Me* biobank.

| ClinVar label | Prediction | Odds ratio | Lower CI | Upper CI | P |
|---|---|---|---|---|---|
| Pathogenic | Recessive | 4.30 | 4.07 | 4.55 | $1.2 \times 10^{-271}$ |
| | Non-Recessive | 1.07 | 1.04 | 1.09 | $2.2 \times 10^{-308}$ |
| Benign | Recessive | 0.98 | 0.97 | 0.98 | $2.2 \times 10^{-308}$ |
| | Non-Recessive | 1.01 | 1.00 | 1.01 | $2.2 \times 10^{-308}$ |
| Uncertain significance | Recessive | 5.45 | 5.13 | 5.77 | $8.4 \times 10^{-250}$ |
| | Non-Recessive | 0.31 | 0.30 | 0.32 | $2.2 \times 10^{-308}$ |
| Conflicting interpretation | Recessive | 4.11 | 3.94 | 4.28 | $2.2 \times 10^{-308}$ |
| | Non-Recessive | 1.11 | 1.09 | 1.13 | $2.2 \times 10^{-308}$ |

Effect sizes (Odds ratios) and 95% confidence intervals were obtained for individual ancestries using a Cochran-Mantel-Haenszel (CMH) test. The reported effect sizes correspond to an inverse variance meta-analysis across ancestries.

**Supplementary Table 6.** Detailed P values and effect sizes for the disease association test on variants predicted Dominant in the Bio*Me* biobank.

| ClinVar label | Prediction | Odds ratio | Lower CI | Upper CI | P |
|---|---|---|---|---|---|
| Pathogenic | Dominant | 1.98 | 1.96 | 2.00 | $2.2 \times 10^{-308}$ |
| | Non-Dominant | 1.56 | 1.55 | 1.57 | $2.2 \times 10^{-308}$ |
| Benign | Dominant | 0.96 | 0.96 | 0.96 | $2.2 \times 10^{-308}$ |
| | Non-Dominant | 1.00 | 1.00 | 1.01 | $2.2 \times 10^{-308}$ |
| Uncertain significance | Dominant | 1.40 | 1.39 | 1.41 | $2.2 \times 10^{-308}$ |
| | Non-Dominant | 0.87 | 0.87 | 0.87 | $2.2 \times 10^{-308}$ |
| Conflicting interpretation | Dominant | 1.13 | 1.12 | 1.13 | $2.2 \times 10^{-308}$ |
| | Non-Dominant | 1.09 | 1.09 | 1.09 | $2.2 \times 10^{-308}$ |

Effect sizes (Odds ratios) and 95% confidence intervals were obtained for individual ancestries using a Cochran-Mantel-Haenszel (CMH) test. The reported effect sizes correspond to an inverse variance meta-analysis across ancestries.

**Supplementary Table 7.** Detailed P values and effect sizes for the disease association test on variants predicted Benign in the Bio*Me* biobank.

| ClinVar label | Prediction | Odds ratio | Lower CI | Upper CI | P |
|---|---|---|---|---|---|
| Pathogenic | Benign | 1.23 | 1.22 | 1.24 | $2.2 \times 10^{-308}$ |
| | Non-Benign | 2.97 | 2.94 | 2.99 | $2.2 \times 10^{-308}$ |
| Benign | Benign | 1.01 | 1.01 | 1.02 | $2.2 \times 10^{-308}$ |
| | Non-Benign | 0.99 | 0.99 | 0.99 | $2.2 \times 10^{-308}$ |
| Uncertain significance | Benign | 0.87 | 0.87 | 0.88 | $2.2 \times 10^{-308}$ |
| | Non-Benign | 1.02 | 1.01 | 1.02 | $2.2 \times 10^{-308}$ |
| Conflicting interpretation | Benign | 1.05 | 1.05 | 1.05 | $2.2 \times 10^{-308}$ |
| | Non-Benign | 1.10 | 1.10 | 1.11 | $2.2 \times 10^{-308}$ |

Effect sizes (Odds ratios) and 95% confidence intervals were obtained for individual ancestries using a Cochran-Mantel-Haenszel (CMH) test. The reported effect sizes correspond to an inverse variance meta-analysis across ancestries.

**Supplementary Table 8.** Detailed P values and effect sizes for the disease association test on variants predicted Recessive in the Bio*Me* biobank, restricted to 2-star review status or higher.

| ClinVar label | Prediction | Odds ratio | Lower CI | Upper CI | P |
|---|---|---|---|---|---|
| Pathogenic | Recessive | 4.48 | 4.16 | 4.80 | $1.4 \times 10^{-162}$ |
| | Non-Recessive | 1.47 | 1.36 | 1.57 | $1.9 \times 10^{-170}$ |
| Benign | Recessive | 0.91 | 0.91 | 0.92 | $2.2 \times 10^{-308}$ |
| | Non-Recessive | 1.01 | 1.01 | 1.01 | $2.2 \times 10^{-308}$ |
| Conflicting interpretation | Recessive | 4.90 | 4.54 | 5.23 | $4.7 \times 10^{-154}$ |
| | Non-Recessive | 3.72 | 3.49 | 3.95 | $1.0 \times 10^{-218}$ |

Effect sizes (Odds ratios) and 95% confidence intervals were obtained for individual ancestries using a Cochran-Mantel-Haenszel (CMH) test. The reported effect sizes correspond to an inverse variance meta-analysis across ancestries.

**Supplementary Table 9.** Detailed P values and effect sizes for the disease association test on variants predicted Dominant in the Bio*Me* biobank, restricted to 2-star review status or higher.

| ClinVar label | Prediction | Odds ratio | Lower CI | Upper CI | P |
|---|---|---|---|---|---|
| Pathogenic | Dominant | 4.18 | 4.13 | 4.22 | $2.2 \times 10^{-308}$ |
| | Non-Dominant | 2.12 | 2.10 | 2.14 | $2.2 \times 10^{-308}$ |
| Benign | Dominant | 0.95 | 0.94 | 0.95 | $2.2 \times 10^{-308}$ |
| | Non-Dominant | 1.01 | 1.01 | 1.01 | $2.2 \times 10^{-308}$ |
| Uncertain significance | Dominant | 1.10 | 1.08 | 1.11 | $2.2 \times 10^{-308}$ |
| | Non-Dominant | 0.90 | 0.90 | 0.91 | $2.2 \times 10^{-308}$ |

Effect sizes (Odds ratios) and 95% confidence intervals were obtained for individual ancestries using a Cochran-Mantel-Haenszel (CMH) test. The reported effect sizes correspond to an inverse variance meta-analysis across ancestries.

**Supplementary Table 10.** Detailed P values and effect sizes for the disease association test on variants predicted Benign in the Bio*Me* biobank, restricted to 2-star review status or higher.

| ClinVar label | Prediction | Odds ratio | Lower CI | Upper CI | P |
|---|---|---|---|---|---|
| Pathogenic | Benign | 1.75 | 1.73 | 1.77 | $2.2 \times 10^{-308}$ |
| | Non-Benign | 4.16 | 4.13 | 4.20 | $2.2 \times 10^{-308}$ |
| Benign | Benign | 1.02 | 1.02 | 1.02 | $2.2 \times 10^{-308}$ |
| | Non-Benign | 0.95 | 0.95 | 0.95 | $2.2 \times 10^{-308}$ |
| Uncertain significance | Benign | 0.93 | 0.92 | 0.94 | $2.2 \times 10^{-308}$ |
| | Non-Benign | 1.00 | 0.98 | 1.01 | $2.2 \times 10^{-308}$ |

Effect sizes (Odds ratios) and 95% confidence intervals were obtained for individual ancestries using a Cochran-Mantel-Haenszel (CMH) test. The reported effect sizes correspond to an inverse variance meta-analysis across ancestries.

**Supplementary Table 11.** Detailed P values and effect sizes for the disease association test on variants predicted Recessive by MAPPIN in the Bio*Me* biobank.

| ClinVar label | Prediction | Odds ratio | Lower CI | Upper CI | P |
|---|---|---|---|---|---|
| Pathogenic | Recessive | 3.61 | 3.41 | 3.80 | $1.6 \times 10^{-297}$ |
| | Non-Recessive | 1.07 | 1.04 | 1.09 | $2.2 \times 10^{-308}$ |
| Benign | Recessive | 1.02 | 1.02 | 1.02 | $2.2 \times 10^{-308}$ |
| | Non-Recessive | 0.97 | 0.97 | 0.98 | $2.2 \times 10^{-308}$ |
| Uncertain significance | Recessive | 0.35 | 0.33 | 0.36 | $2.2 \times 10^{-308}$ |
| | Non-Recessive | 0.25 | 0.24 | 0.26 | $2.2 \times 10^{-308}$ |
| Conflicting interpretation | Recessive | 1.15 | 1.11 | 1.18 | $2.2 \times 10^{-308}$ |
| | Non-Recessive | 1.15 | 1.13 | 1.17 | $2.2 \times 10^{-308}$ |

Effect sizes (Odds ratios) and 95% confidence intervals were obtained for individual ancestries using a Cochran-Mantel-Haenszel (CMH) test. The reported effect sizes correspond to an inverse variance meta-analysis across ancestries.

**Supplementary Table 12.** Detailed P values and effect sizes for the disease association test on variants predicted Dominant by MAPPIN in the Bio*Me* biobank.

| ClinVar label | Prediction | Odds ratio | Lower CI | Upper CI | P |
|---|---|---|---|---|---|
| Pathogenic | Dominant | 8.92 | 8.83 | 9.00 | $2.2 \times 10^{-308}$ |
| | Non-Dominant | 1.48 | 1.47 | 1.49 | $2.2 \times 10^{-308}$ |
| Benign | Dominant | 1.04 | 1.04 | 1.04 | $2.2 \times 10^{-308}$ |
| | Non-Dominant | 1.03 | 1.03 | 1.03 | $2.2 \times 10^{-308}$ |
| Uncertain significance | Dominant | 1.02 | 1.01 | 1.03 | $2.2 \times 10^{-308}$ |
| | Non-Dominant | 0.83 | 0.83 | 0.83 | $2.2 \times 10^{-308}$ |
| Conflicting interpretation | Dominant | 1.15 | 1.15 | 1.16 | $2.2 \times 10^{-308}$ |
| | Non-Dominant | 0.97 | 0.97 | 0.97 | $2.2 \times 10^{-308}$ |

Effect sizes (Odds ratios) and 95% confidence intervals were obtained for individual ancestries using a Cochran-Mantel-Haenszel (CMH) test. The reported effect sizes correspond to an inverse variance meta-analysis across ancestries.

**Supplementary Table 13.** Detailed P values and effect sizes for the disease association test on variants predicted Benign by MAPPIN in the Bio*Me* biobank.

| ClinVar label | Prediction | Odds ratio | Lower CI | Upper CI | P |
|---|---|---|---|---|---|
| Pathogenic | Benign | 1.71 | 1.70 | 1.71 | $2.2 \times 10^{-308}$ |
| | Non-Benign | 3.21 | 3.19 | 3.23 | $2.2 \times 10^{-308}$ |
| Benign | Benign | 1.03 | 1.03 | 1.04 | $2.2 \times 10^{-308}$ |
| | Non-Benign | 1.01 | 1.01 | 1.01 | $2.2 \times 10^{-308}$ |
| Uncertain significance | Benign | 0.59 | 0.59 | 0.60 | $2.2 \times 10^{-308}$ |
| | Non-Benign | 0.93 | 0.92 | 0.93 | $2.2 \times 10^{-308}$ |
| Conflicting interpretation | Benign | 0.90 | 0.90 | 0.90 | $2.2 \times 10^{-308}$ |
| | Non-Benign | 1.18 | 1.18 | 1.18 | $2.2 \times 10^{-308}$ |

Effect sizes (Odds ratios) and 95% confidence intervals were obtained for individual ancestries using a Cochran-Mantel-Haenszel (CMH) test. The reported effect sizes correspond to an inverse variance meta-analysis across ancestries.

**Supplementary Table 14.** Description of significant SNP associations with disease on African Americans.

| SNP ID | Gene | AF | P value | OR (95% CI) |
|---|---|---|---|---|
| rs79985808 | SUMF1 | 0.041 | $9.13 \times 10^{-3}$ | 65.36 (2.82 to 1,522) |
| rs17144835 | DNAH11 | 0.075 | 0.10 | 9.26 (0.63 to 135.87) |
| rs1800562 | HFE | 0.011 | 0.53 | 4.31 (0.04 to 407.3) |
| rs145214720 | COL10A1 | $3.34 \times 10^{-4}$ | 0.091 | 8.65 (0.71 to 105.22) |
| rs34539681 | COL10A1 | 0.010 | $1.49 \times 10^{-3}$ | 1,406.10 (16.05 to 123,123) |
| rs140075817 | EXT2 | $8.35 \times 10^{-5}$ | 0.074 | 127.32 (0.62 to 26,007) |
| rs770821909 | EXT2 | $3.34 \times 10^{-4}$ | $5.91 \times 10^{-4}$ | 911.04 (18.67 to 44,462) |
| rs146098187 | EXT2 | 0.00 | NA | NA |
| rs138495222 | EXT2 | $8.35 \times 10^{-5}$ | 0.19 | 48.25 (0.14 to 16,001) |
| rs35221558 | LEMD3 | $5.84 \times 10^{-4}$ | $2.09 \times 10^{-3}$ | 155.68 (6.24 to 3,883) |
| rs36105360 | LMNB1 | $4.26 \times 10^{-3}$ | 0.12 | 6.67 (0.61 to 73.36) |
| rs139644798 | RARS1 | $1.67 \times 10^{-4}$ | 0.015 | 117.58 (2.51 to 549.00) |
| rs34637584 | LRRK2 | $8.35 \times 10^{-5}$ | 0.061 | 185.17 (2.51 to 4,402) |
| rs141230910 | SDHB | $2.20 \times 10^{-3}$ | 0.012 | 8.38 (1.58 to 44.39) |
| rs372115732 | TBX4 | $8.35 \times 10^{-5}$ | $3.05 \times 10^{-4}$ | 69,390 (163.45 to 29,458,080) |
| rs141707850 | FBN2 | $8.35 \times 10^{-5}$ | 0.029 | 180.95 (1.71 to 1,908) |
| rs147272790 | MBD5 | $7.51 \times 10^{-4}$ | $3.32 \times 10^{-6}$ | 23.09 (6.15 to 86.76) |
| rs77375493 | JAK2 | $3.34 \times 10^{-4}$ | $9.08 \times 10^{-3}$ | 15.22 (1.96 to 117.64) |

The significance threshold is set to $p=1.09 \times 10^{-4}$ for the recessive association test and $p=7.83 \times 10^{-6}$ for the dominant association test after a Bonferroni correction based on 455 and 6,382 tests respectively. AF corresponds to allele frequency, OR corresponds to odds ratio, CI corresponds to confidence interval.

**Supplementary Table 15.** Description of significant SNP associations with disease on European Americans.

| SNP ID | Gene | AF | P value | OR (95% CI) |
|---|---|---|---|---|
| rs79985808 | SUMF1 | $1.40 \times 10^{-4}$ | NA | NA |
| rs17144835 | DNAH11 | 0.071 | $2.39 \times 10^{-3}$ | 54.33 (3.70 to 797.46) |
| rs1800562 | HFE | 0.042 | $1.18 \times 10^{-9}$ | 14.37 (6.09 to 33.94) |
| rs145214720 | COL10A1 | $4.21 \times 10^{-4}$ | 0.15 | 6.07 (0.51 to 71.79) |
| rs34539681 | COL10A1 | $1.40 \times 10^{-4}$ | $7.41 \times 10^{-3}$ | 185.17 (4.05 to 8,456) |
| rs140075817 | EXT2 | $3.35 \times 10^{-4}$ | $5.81 \times 10^{-3}$ | 142.90 (4.20 to 4,858) |
| rs770821909 | EXT2 | 0.00 | NA | NA |
| rs146098187 | EXT2 | $9.13 \times 10^{-4}$ | $3.70 \times 10^{-4}$ | 342.18 (13.78 to 8,496) |
| rs138495222 | EXT2 | $8.43 \times 10^{-4}$ | $2.21 \times 10^{-3}$ | 103.71 (5.30 to 2,027) |
| rs35221558 | LEMD3 | $4.00 \times 10^{-3}$ | 0.028 | 22.14 (1.40 to 348.80) |
| rs36105360 | LMNB1 | 0.014 | $5.21 \times 10^{-3}$ | 6.43 (1.74 to 23.73) |
| rs139644798 | RARS1 | $4.90 \times 10^{-4}$ | 0.29 | 31.42 (1.43 to 688.60) |
| rs34637584 | LRRK2 | $3.16 \times 10^{-3}$ | $2.19 \times 10^{-5}$ | 4.97 (2.37 to 10.43) |
| rs141230910 | SDHB | $1.40 \times 10^{-4}$ | 0.18 | 13.34 (0.31 to 567.71) |
| rs372115732 | TBX4 | $2.11 \times 10^{-4}$ | 0.035 | 43.70 (1.29 to 1,473) |
| rs141707850 | FBN2 | $3.51 \times 10^{-4}$ | 0.029 | 42.57 (1.46 to 1,241.08) |
| rs147272790 | MBD5 | 0.00 | NA | NA |
| rs77375493 | JAK2 | $9.13 \times 10^{-4}$ | $1.93 \times 10^{-7}$ | 20.22 (6.52 to 62.72) |

The significance threshold is set to $p=1.09 \times 10^{-4}$ for the recessive association test and $p=7.83 \times 10^{-6}$ for the dominant association test after a Bonferroni correction based on 455 and 6,382 tests respectively. AF corresponds to allele frequency, OR corresponds to odds ratio, CI corresponds to confidence interval.

**Supplementary Table 16.** Description of significant SNP associations with disease on Hispanic Americans.

| SNP ID | Gene | AF | P value | OR (95% CI) |
|---|---|---|---|---|
| rs79985808 | SUMF1 | 0.014 | 0.014 | 171.40 (2.74 to 10,712) |
| rs17144835 | DNAH11 | 0.052 | $7.0 \times 10^{-3}$ | 36.39 (2.56 to 517.37) |
| rs1800562 | HFE | 0.015 | NA | NA |
| rs145214720 | COL10A1 | $2.49 \times 10^{-4}$ | 0.21 | 4.95 (0.41 to 58.96) |
| rs34539681 | COL10A1 | $3.54 \times 10^{-3}$ | $2.04 \times 10^{-3}$ | 34,986 (45.25 to 27,045,040) |
| rs140075817 | EXT2 | $3.11 \times 10^{-4}$ | $5.78 \times 10^{-4}$ | 616.34 (15.88 to 23,914) |
| rs770821909 | EXT2 | $6.22 \times 10^{-5}$ | 0.019 | 275.77 (2.53 to 30,069) |
| rs146098187 | EXT2 | $6.22 \times 10^{-5}$ | 0.022 | 256.97 (2.22 to 29,675) |
| rs138495222 | EXT2 | $1.24 \times 10^{-4}$ | $8.88 \times 10^{-3}$ | 214.07 (3.84 to 11,919) |
| rs35221558 | LEMD3 | $1.31 \times 10^{-3}$ | 0.012 | 32.17 (2.13 to 484.55) |
| rs36105360 | LMNB1 | $9.40 \times 10^{-3}$ | 0.018 | 6.32 (1.38 to 29.07) |
| rs139644798 | RARS1 | $6.22 \times 10^{-5}$ | 0.023 | 1,199.72 (2.66 to 540,262) |
| rs34637584 | LRRK2 | $2.24 \times 10^{-3}$ | 0.12 | 2.83 (0.76 to 10.46) |
| rs141230910 | SDHB | $3.11 \times 10^{-4}$ | $1.25 \times 10^{-4}$ | 66.32 (7.77 to 565.93) |
| rs372115732 | TBX4 | $6.22 \times 10^{-5}$ | 0.047 | 105.75 (1.05 to 1,062.13) |
| rs141707850 | FBN2 | $6.22 \times 10^{-5}$ | $4.48 \times 10^{-3}$ | 1,581.87 (9.83 to 254,494) |
| rs147272790 | MBD5 | $1.24 \times 10^{-4}$ | 0.41 | 4.78 (0.11 to 198.34) |
| rs77375493 | JAK2 | $4.35 \times 10^{-4}$ | $1.66 \times 10^{-4}$ | 17.75 (3.97 to 79.35) |

The significance threshold is set to $p=1.09 \times 10^{-4}$ for the recessive association test and $p=7.83 \times 10^{-6}$ for the dominant association test after a Bonferroni correction based on 455 and 6,382 tests respectively. AF corresponds to allele frequency, OR corresponds to odds ratio, CI corresponds to confidence interval.

**Supplementary Table 17.** Description of significant SNP associations with disease on other ancestries.

| SNP ID | Gene | AF | P value | OR (95% CI) |
|---|---|---|---|---|
| rs79985808 | SUMF1 | 5.97 x $10^{-3}$ | 0.31 | 752 (1.81 to 312,365) |
| rs17144835 | DNAH11 | 0.038 | 3.53 x $10^{-3}$ | 142.18 (5.08 to 3,975) |
| rs1800562 | HFE | 0.011 | NA | NA |
| rs145214720 | COL10A1 | 1.66 x $10^{-4}$ | 0.19 | 4.87 (0.45 to 52.39) |
| rs34539681 | COL10A1 | 6.64 x $10^{-4}$ | 2.08 x $10^{-3}$ | 227.66 (7.18 to 7,214) |
| rs140075817 | EXT2 | 0.00 | NA | NA |
| rs770821909 | EXT2 | 1.66 x $10^{-4}$ | 0.023 | 287.26 (2.18 to 40,518) |
| rs146098187 | EXT2 | 1.66 x $10^{-4}$ | 0.049 | 143.84 (2.18 to 20,372) |
| rs138495222 | EXT2 | 4.98 x $10^{-4}$ | 5.64 x $10^{-3}$ | 691.97 (6.74 to 71,022) |
| rs35221558 | LEMD3 | 9.96 x $10^{-4}$ | 0.015 | 34.17 (1.97 to 593.61) |
| rs36105360 | LMNB1 | 0.015 | 2.31 x $10^{-3}$ | 14.79 (2.61 to 83.69) |
| rs139644798 | RARS1 | 8.30 x $10^{-4}$ | 5.73 x $10^{-3}$ | 166.41 (4.42 to 6,263) |
| rs34637584 | LRRK2 | 1.66 x $10^{-4}$ | 0.16 | 28.60 ( 0.26 to 3,140) |
| rs141230910 | SDHB | 1.66 x $10^{-4}$ | 0.14 | 34.53 ( 0.30 to 3,897) |
| rs372115732 | TBX4 | 1.66 x $10^{-4}$ | 7.39 x $10^{-3}$ | 290.43 (4.57 to 18,420) |
| rs141707850 | FBN2 | 1.66 x $10^{-4}$ | 7.56 x $10^{-3}$ | 190.73 (4.03 to 9,011) |
| rs147272790 | MBD5 | 1.66 x $10^{-4}$ | 0.21 | 18.36 (0.19 to 1,786) |
| rs77375493 | JAK2 | 4.93 x $10^{-4}$ | 0.029 | 24,49 (1.37 to 436.98) |

The significance threshold is set to p=1.09 x $10^{-4}$ for the recessive association test and p=7.83 x $10^{-6}$ for the dominant association test after a Bonferroni correction based on 455 and 6,382 tests respectively. AF corresponds to allele frequency, OR corresponds to odds ratio, CI corresponds to confidence interval.
